# Long-Term Impact of Molecular Epidemiology Shifts of Methicillin-resistant *Staphylococcus aureus* on Severity and Mortality of Bloodstream Infection

**DOI:** 10.1101/2024.02.15.24302553

**Authors:** Norihito Kaku, Masaki Ishige, Go Yasutake, Daisuke Sasaki, Kenji Ota, Fujiko Mitsumoto-Kaseida, Kosuke Kosai, Hiroo Hasegawa, Koichi Izumikawa, Hiroshi Mukae, Katsunori Yanagihara

**Author notes:** **Corresponding Author:** Norihito Kaku 1-7-1 Sakamoto, Nagasaki City, Nagasaki, Japan 852-8501.

## Abstract

A 2019 nationwide study in Japan revealed the predominant methicillin-resistant Staphylococcus aureus (MRSA) types in bloodstream infections (BSIs) to be sequence type (ST)8-carrying SCC*mec* type IV (ST8-MRSA-IV) and clonal complex 1-carrying SCC*mec* type IV (CC1- MRSA-IV). However, detailed patient characteristics and how these MRSA types evolve over time remain largely unknown. In this long-term single-center study, MRSA strains isolated from blood cultures at Nagasaki University Hospital from 2012 to 2019 were sequenced and analyzed. Additionally, we compared the SCC*mec* types and patient characteristics identified in this study with previous data from our hospital spanning 2003 to 2007 and 2008 to 2011. Over this 16-year period, SCC*mec* type II decreased significantly from 79.2% to 15.5%, while type IV increased from 18.2% to 65.5%. This shift in SCC*mec* types was associated with notable changes in severity and outcomes; the sequential organ failure assessment (SOFA) score decreased from 5.8 to 3.1; in-hospital mortality declined from 39.8% to 15.5%. In contrast, no significant changes in patient demographics, such as age, sex, or underlying diseases, were observed. Between 2012 and 2019, the major combinations of SCC*mec* type and sequence type were ST8-MRSA-IV, ST8-MRSA-I, CC1-MRSA-IV, and ST5-MRSA-II. Additionally, ST8-MRSA-IV was divided into CA-MRSA/J, t5071-ST8-MRSA-IV, and USA300-like clone based on the results of molecular analysis. These major combinations showed similar drug resistance patterns, molecular characteristics, and phylogenetic features to those identified in nationwide surveillance. This study highlights the evolving nature of MRSA types in bloodstream infections, correlating with improved patient outcomes over time.

## Introduction

Bloodstream infection (BSI) is a severe condition that can lead to sepsis, a systemic inflammatory response. Therefore, administering proper treatment as early as possible is crucial. However, when drug-resistant bacteria cause BSI, appropriate treatment may be delayed. Methicillin-resistant *Staphylococcus aureus* (MRSA) is the most common causative multidrug-resistant pathogen of BSI.^1,2^ In Japan and the United States, the predominant MRSA strain was the New York/Japan clone, which is defined as ST5 harboring the SCC*mec* II island (ST5- MRSA-II) and exhibiting multidrug resistance.^3^ However, our previous report revealed a decrease in SCC*mec* type II from 79.2% between 2003 and 2007 to 44.9% between 2008 and 2011; conversely, SCC*mec* types I and IV increased.^4,5^ According to nationwide surveillance conducted in 2019, ST8 carrying SCC*mec* type IV (ST8-MRSA-IV) and clonal complex 1 carrying SCC*mec* type IV (CC1-MRSA-IV) were the predominant molecular types in BSI.^6^ Additionally, ST8-MRSA-IV is subdivided into several groups based on molecular characteristics.^6^ However, owing to the limited number of MRSA strains detected at each facility during the surveillance, it remains undetermined whether similar trends are observed in individual facilities. Furthermore, the differences in the backgrounds of patients infected with various MRSA types remain unknown.

Therefore, in this study, we aimed to achieve three main objectives. First, by incorporating the results of our two previous reports,^4,5^ we aimed to investigate the long-term changes in SCC*mec* types, as well as changes in patient characteristics and clinical outcomes from 2003 to 2019. Second, using MRSA strains isolated from 2012 to 2019, we performed whole-genome sequencing (WGS) to analyze the molecular epidemiological characteristics of MRSA detected in BSIs at our hospital since 2012 and to assess how these changes align with the trends observed in the 2019 nationwide surveillance. Additionally, we investigated the difference in patient background, clinical outcomes, and positive rate for drug-resistance genes and virulence genes among the major MRSA types.

## Materials and methods

### Study design

This retrospective observational study was conducted at Nagasaki University Hospital, Nagasaki, Japan, from January 2012 to December 2019.^5^ Nagasaki University Hospital is a tertiary medical institution with 874 beds in Nagasaki Prefecture, the westernmost part of Japan. The number of MRSA detected from blood cultures per 1,000 patient days was not changed between 2003 and 2019 (0.148 and 0.145, respectively), while the number of methicillin-susceptance *Staphylococcus aureus* (MSSA) per 1,000 patient days increased from 0.042 in 2003 to 0.257 in 2019 (Supplementary Fig. 1). In Nagasaki University Hospital, screening and decolonization for MRSA were performed at the discretion of each the Intensive Care Unit (ICU), Neonatal Intensive Care Unit (NICU)/General Care Unit (GCU), Orthopedics, and Cardiovascular Surgery. The infection control team did not proactively intervene in MRSA treatment; instead, it intervened only upon consultation by the principal physicians. No special infection control measures were implemented solely based on the detection of MRSA. However, if an outbreak was suspected, the infection control team investigated to identify the source and implemented appropriate measures.

Patient data, including blood culture collection location (outpatient or inpatient), the day of hospitalization blood culture collection, antimicrobial use during the 30 days before the onset of BSI, initial antimicrobial regimens for BSI, mortality, and the details required to calculate the Charlson comorbidity index^7^ and Sequential Organ Failure Assessment (SOFA) score^8^ were collected from the medical records. MRSA BSI was classified as community-acquired, healthcare-associated, or hospital-acquired following the previous report.^9^ MRSA strains isolated from blood cultures from 2012 to 2019 were subjected to antimicrobial susceptibility testing (AST) and whole-genome sequencing (WGS). To evaluate long-term trends in SCC*mec* types, patient characteristics, and clinical outcomes, data from 2003 to 2011 were retrieved from the databases used in our previous studies.^4,5^ To match the study period of previous studies with that of this study, the data obtained in this study was analyzed separately for four years, 2012-2015 and 2016-2019.

### Strain collection

In the previous studies conducted between January 2003 and December 2011,^4,5,^ MRSA strains detected in one or more blood cultures were analyzed because collecting more than two sets of blood cultures was uncommon at our hospital. However, collecting two or more sets of blood cultures has become more common since the 2010s. Therefore, in this study, we analyzed MRSA strains detected in two or more blood cultures between January 2012 and December 2019 using the same inclusion criteria for nationwide surveillance.^6^ To avoid redundancy, only the first isolate identified was included in the analysis when multiple isolates were detected from the same patient during a single hospitalization.

### Antimicrobial susceptibility testing

We measured the minimum inhibitory concentrations (MICs) using broth microdilution testing using a dry plate (Eiken, Tokyo, Japan), according to the manufacturer’s instructions. Antimicrobial susceptibility was determined according to the Clinical and Laboratory Standards Institute guidelines (CLSI M100-Ed31) and the European Committee on Antimicrobial Susceptibility Testing Version 11.0.

### Whole-genome sequencing

All procedures were performed according to the respective manufacturer’s instructions. After extracting the DNA from MRSA strains using a Quick-DNA Fungal/Bacterial Kit (ZYMO RESEARCH, Irvine, CA, United States), we performed WGS on the MRSA strains isolated between 2012 and 2015 using an Ion PGM HiQ View OT2 Kit (Thermo Fisher Scientific, Waltham, MA, United States). Enriched samples were loaded onto an Ion 318 chip and sequenced using an IonTorrent Personal Genome Analyzer with an Ion PGM HiQ View Sequencing Kit (Thermo Fisher Scientific). DNA libraries were generated for Ion PGM using the Ion Xpress Plus Fragment Library Kit (4471269; Thermo Fisher Scientific). We performed WGS for the MRSA strains isolated between 2016 and 2019 using the MiSeq system (Illumina, San Diego, CA, United States) and MiSeq Reagent kit v3 (600 cycles) (Illumina). We also performed WGS for MRSA strains isolated between 2012 and 2015, which needed to be retested using the MiSeq system. We generated DNA libraries for the Miseq system using Invitrogen (Waltham, MA, United States) Collibri ES DNA Library Prep Kit for Illumina (A38607096, ThermoFisher Scientific).

### Analysis of molecular characteristics

Sequence data were assembled using the CLC Genomics Workbench Microbial Genomics Module (Qiagen, Venlo, Netherlands). Multilocus sequence type (MLST) was determined using PubMLST (https://pubmlst.org).^10^ We determined the SCC*mec* and spa types using SCCmecFinder (ver1.2)^11^ and spaTyper (software version 1.0 and database version 2023-6-19)^12^ on the Center for Genomic Epidemiology website (https://www.genomicepidemiology.org), respectively. Resistance and virulence genes were detected using ResFinder (software version 2023-08-22 and database version 2023-04-12) and VirulenceFinder (software version 2.0.3 and database version 2022-12-02)^13,14^ on the Center for Genomic Epidemiology website (https://www.genomicepidemiology.org), respectively. Our previous study revealed very similar bacteriological characteristics between ST1-carrying SCC*mec* type IV (ST1-MRSA-IV) and ST2725-carrying SCC*mec* type IV (ST2725-MRSA-IV);^6^ further, a preliminary investigation as part of the current study showed that ST1-MRSA-IV, ST2725-MRSA-IV, and ST5213-MRSA-IV had the same characteristics. Hence, we combined the data from ST1-MRSA-IV, ST2725- MRSA-IV, and ST5213-MRSA-IV as CC1-MRSA-IV. Core-genome MLST (cgMLST) was performed using Ridom SeqSphere+ v.9.0.10 (Ridom GmbH, Münster, Germany. A minimum spanning tree (MST) or unweighted pair group method with arithmetic mean (UPGMA) tree was created based on MLST, cgMLST, and *S. aureus* accessories using Ridom SeqSphere+ (ver. 9). Samples with more than 10% missing values among the data used for the distance calculation were excluded from constructing the MST or UPGMA. For the UPGMA using strains that were detected during the previous nationwide surveillance,^6^ we excluded 13 strains detected at Nagasaki University Hospital to avoid duplication. In UPGMA, a cluster of MRSA strains was classified with a distance of 0.1, and subclusters of ST8 with SCC*mec* type I (ST8-MRSA-I) and ST8-MRSA-IV, CC1-MRSA-IV, and ST5-MRSA-II were classified with a distance of 0.03.

### Statistical analysis

All statistical analyses except q-value calculation were performed on GraphPad Prism 10 (version 10.1.0; GraphPad Software, Boston, MA, United States of America). Continuous variables were expressed as mean ± standard deviation. Categorical variables were compared using Fisher’s exact test, and q-values were calculated using the Benjamini–Hochberg method for multiple comparisons. Continuous variables were compared using Student’s t-test for two-group comparisons and Tukey’s test for multiple comparisons. The statistical significance level was set at *p* < 0.05 and *q* < 0.05, and the *p*-value was considered not significant (n.s.) if it was greater than 0.2.

### Ethics

This study was approved by the Ethics Committee of the Nagasaki University Hospital (20072018). MRSA strains collected from blood cultures were anonymized and individually numbered. Patient information collected from the medical records was also anonymized and individually numbered when. The Ethics Committee of Nagasaki University Hospital waived the requirement for informed consent.

## Results

### Changes in SCCmec type, patient characteristics, and outcomes from 2003 to 2019

Twenty-seven and fifty-eight MRSA strains were isolated during 2012–2015 and 2016–2019, respectively. The percentages of SCC*mec* types I, II, and IV during 2012–2015 were 37.0%, 37.0%, and 25.9%, respectively. The percentages of SCC*mec* types I, II, IV, and V during 2016– 2019 were 15.5%, 15.5%, 65.5%, and 3.4%, respectively. Fig. 1 and Supplementary Table 1 show the changes in SCC*mec* type from 2003 to 2019. SCC*mec* type II was the most frequently detected type from 2003 to 2015. However, its frequency decreased significantly from 79.2% during 2003–2007 to 15.5% during 2016–2019 (*q* < 0.001). In contrast, the percentage of SCC*mec* type IV increased dramatically from 18.2% during 2003–2007 to 65.5% during 2016– 2019 (*q* < 0.001). SCC*mec* type I showed changes that differed from those of SCC*mec* types II and IV. The percentage of SCC*mec* type I cases increased significantly from 2.6% during 2003–2007 to 37.0% during 2012–2015 (*q* < 0.001). SCC*mec* type I was one of the most frequently detected SCC*mec* types from 2012 to 2015; however, its prevalence decreased by more than 50% (15.5 %) during 2016–2019.Table 1 and Supplementary Table 1 show the characteristics of patients in each study period. There were no significant differences in the patients’ backgrounds, such as age, sex, or underlying diseases, except for the classification of infection; the percentage of healthcare-associated infections was significantly higher during 2012–2015 than during 2008– 2011 (3.6% vs. 22.2%, *q* = 0.020). Further, the sources of MRSA infection changed from 2003 to 2019. The percentage of intravascular device-related BSI increased significantly from 18.1% during 2003–2007 to 46.6% during 2016–2019 (*q* = 0.002). In contrast, the percentage of respiratory tract infection-related BSI significantly decreased from 16.9% during 2003–2007 to 20.5% during 2008–2011 and 1.7% during 2016–2019 (*q* = 0.013 and 0.004, respectively). The severity of BSI also changed between 2003 and 2019. SOFA score significantly decreased from 5.8 ± 0.5 during 2003–2007 to 3.1 ± 0.5 during 2016–2019 (*p* = 0.004), and accordingly, the in-hospital mortality improved from 39.8% during 2003–2007 to 15.5% during 2016–2019 (*q* = 0.015). As in the previous studies,^4,5^ when MRSA strains detected in one or more blood cultures were included, the in-hospital mortality was 24.3% (17/70) during 2012-2015 and 17.0% (16/94) during 2016-2019, with a significantly lower mortality rate during 2016-2019 compared to 2003- 2007 (*q* = 0.008).

**Fig. 1.**
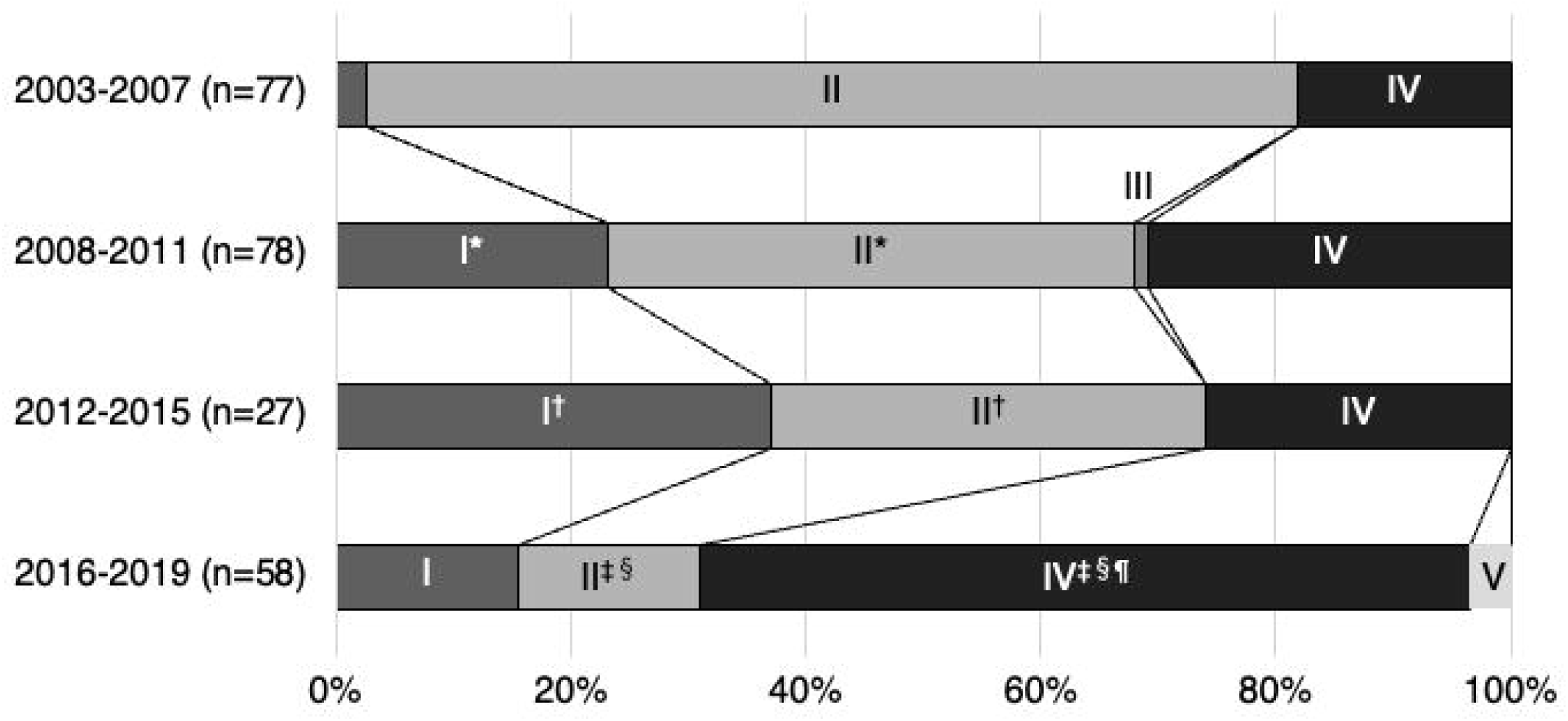
Changes in SCC*mec* type from 2003 to 2019. Changes in SCC*mec* type from 2003 to 2019 were evaluated. Data regarding SCC*mec* type during 2003–2007 and 2008–2011 have been reported previously. The inclusion criteria for MRSA strains in this study (2012–2015 and 2016–2019) differed from those used in previous studies (2003–2007 and 2008–2011). In previous studies, MRSA strains collected from one or more blood cultures were analyzed, whereas in this study, MRSA strains collected from two or more blood cultures were analyzed. Statistically significant differences were observed in *2003- 2007 versus 2008-2011, ^†^2003-2007 versus 2012-2015, ^‡^2003-2007 versus 2016-2019, ^§^2008-2011 versus 2016-2019, and ^¶^ 2012-2015 versus 2016-2019.

**Table 1.**
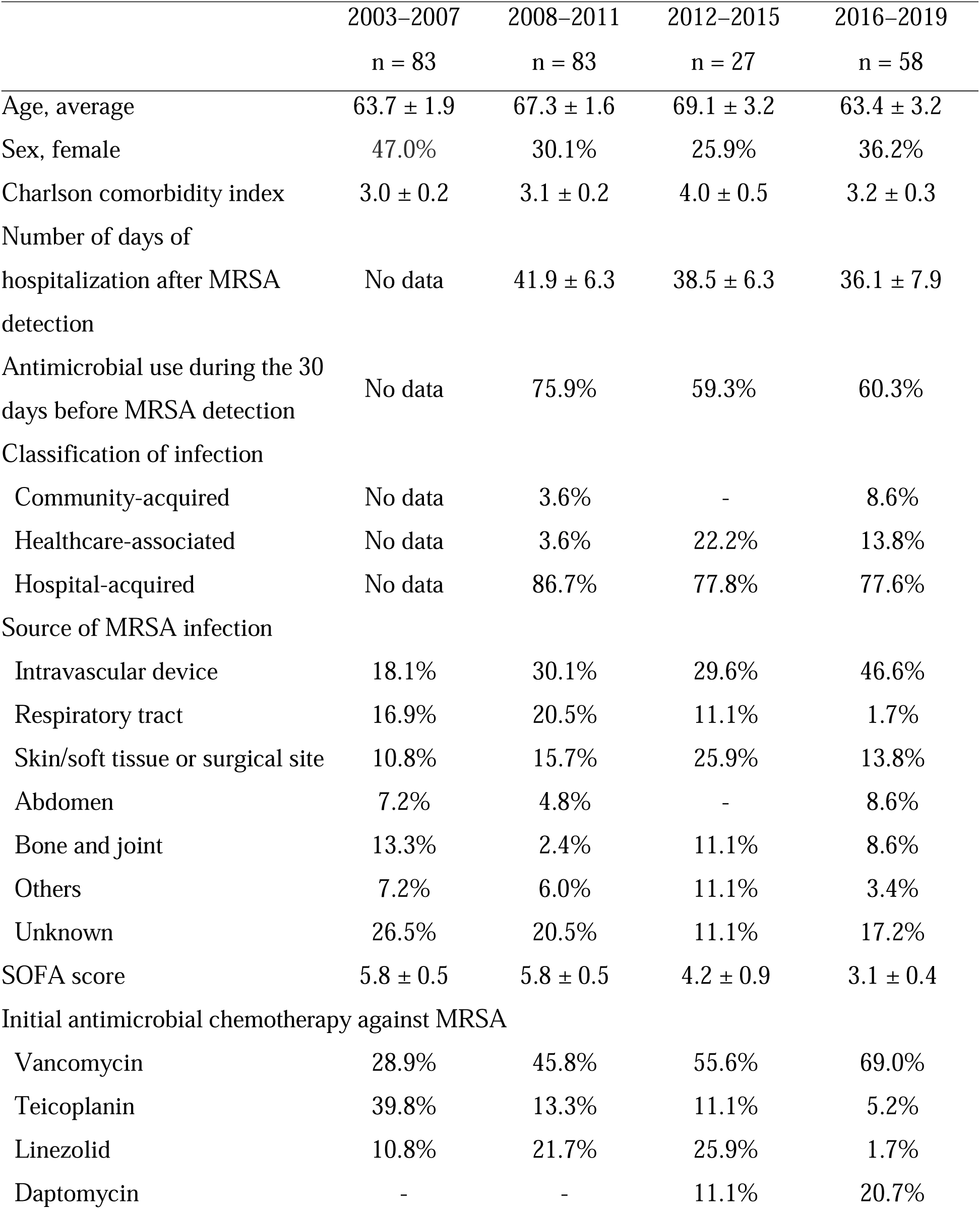
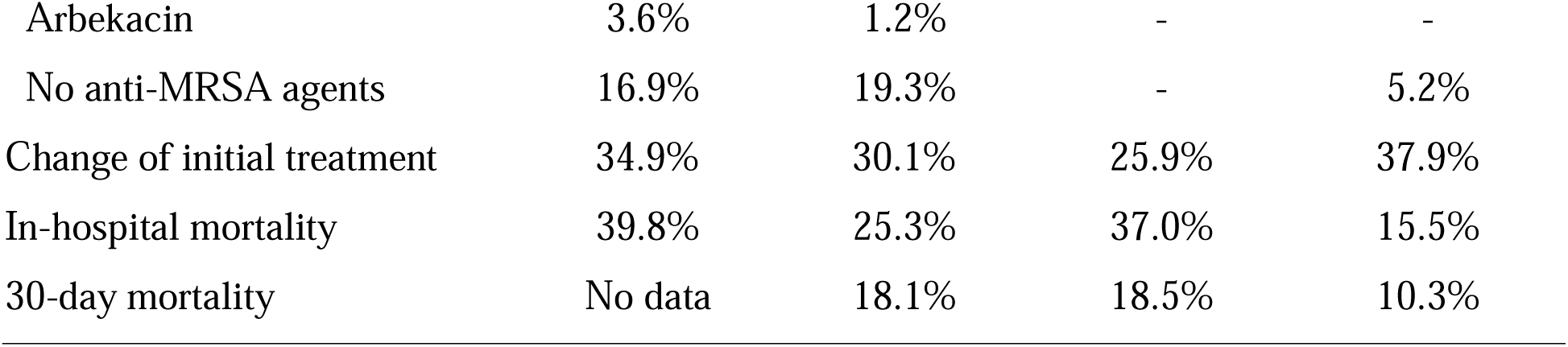
Changes in patient characteristics from 2003 to 2019.

|  | 2003–2007<br>n = 83 | 2008–2011<br>n = 83 | 2012–2015<br>n = 27 | 2016–2019<br>n = 58 |
| --- | --- | --- | --- | --- |
| Age, average | 63.7 ± 1.9 | 67.3 ± 1.6 | 69.1 ± 3.2 | 63.4 ± 3.2 |
| Sex, female | 47.0% | 30.1% | 25.9% | 36.2% |
| Charlson comorbidity index | 3.0 ± 0.2 | 3.1 ± 0.2 | 4.0 ± 0.5 | 3.2 ± 0.3 |
| Number of days of hospitalization after MRSA detection | No data | 41.9 ± 6.3 | 38.5 ± 6.3 | 36.1 ± 7.9 |
| Antimicrobial use during the 30 days before MRSA detection | No data | 75.9% | 59.3% | 60.3% |
| Classification of infection |  |  |  |  |
| Community-acquired | No data | 3.6% | - | 8.6% |
| Healthcare-associated | No data | 3.6% | 22.2% | 13.8% |
| Hospital-acquired | No data | 86.7% | 77.8% | 77.6% |
| Source of MRSA infection |  |  |  |  |
| Intravascular device | 18.1% | 30.1% | 29.6% | 46.6% |
| Respiratory tract | 16.9% | 20.5% | 11.1% | 1.7% |
| Skin/soft tissue or surgical site | 10.8% | 15.7% | 25.9% | 13.8% |
| Abdomen | 7.2% | 4.8% | - | 8.6% |
| Bone and joint | 13.3% | 2.4% | 11.1% | 8.6% |
| Others | 7.2% | 6.0% | 11.1% | 3.4% |
| Unknown | 26.5% | 20.5% | 11.1% | 17.2% |
| SOFA score | 5.8 ± 0.5 | 5.8 ± 0.5 | 4.2 ± 0.9 | 3.1 ± 0.4 |
| Initial antimicrobial chemotherapy against MRSA |  |  |  |  |
| Vancomycin | 28.9% | 45.8% | 55.6% | 69.0% |
| Teicoplanin | 39.8% | 13.3% | 11.1% | 5.2% |
| Linezolid | 10.8% | 21.7% | 25.9% | 1.7% |
| Daptomycin | - | - | 11.1% | 20.7% |
| Arbekacin | 3.6% | 1.2% | - | - |
| No anti-MRSA agents | 16.9% | 19.3% | - | 5.2% |
| Change of initial treatment | 34.9% | 30.1% | 25.9% | 37.9% |
| In-hospital mortality | 39.8% | 25.3% | 37.0% | 15.5% |
| 30-day mortality | No data | 18.1% | 18.5% | 10.3% |

### MRSA types based on ST and SCCmec type from 2012 to 2019

The predominant combination of ST and SCC*mec* types during 2012–2019 was ST8-MRSA-IV (37.6%), followed by ST8-MRSA-I (22.4%), ST5-MRSA-II) (18.8%), and CC1-MRSA-IV (9.4%) (Fig. 2A). The changes in the major combinations are shown in Fig. 2B and Supplementary Table 2. Although there was no change in the percentage of ST8 between 2012– 2015 and 2016–2019, the combination of SCC*mec* type with ST8 differed between the two periods. The percentage of ST8-MRSA-IV was higher during 2016–2019 (44.8%) than during 2012–2015 (22.2%; *p* = 0.056). In contrast, the percentage of ST8-MRSA-I was significantly lower during 2016–2019 (15.5%) than during 2012–2015 (37.0%; *p* = 0.048).

**Fig. 2.**
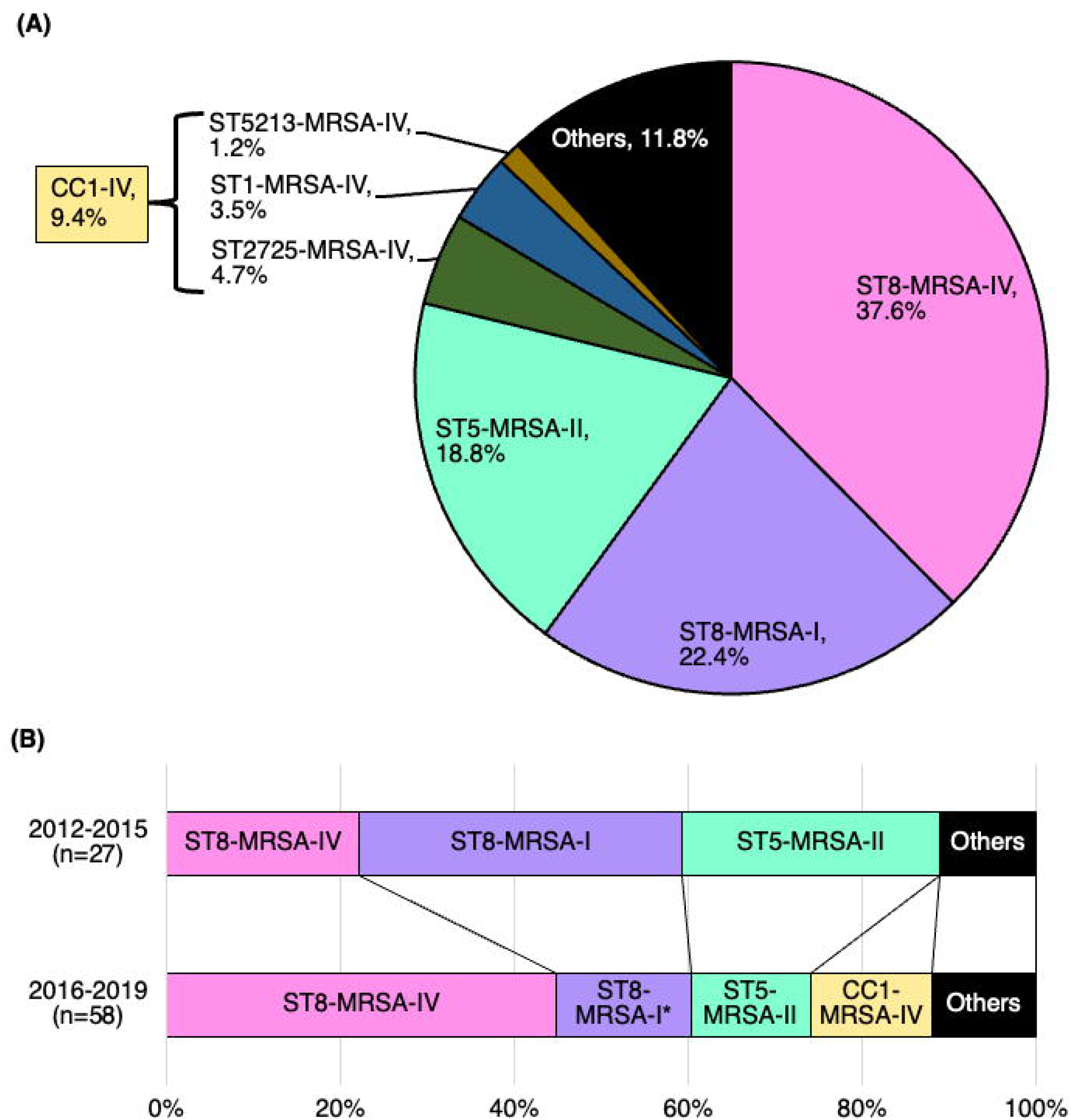
Major MRSA types based on sequence type and SCC*mec* type from 2012 to 2019. SCC*mec* type and sequence type (ST) from 2012 to 2019 were evaluated (A). The changes in the percentage of major MRSA types based on the SCC*mec* type and ST from 2012 to 2019 were also evaluated (**B**). We decided on ST based on whole-genome sequencing using the CLC Genomics Workbench and the CLC Microbial Genomics Module (Qiagen, Venlo, Netherlands).

### Patient characteristics and drug-resistance rate in the major MRSA types

Table 2 and Supplementary Table 3 show the patient characteristics for each major combination. No significant differences were observed with respect to the patient characteristics among the major MRSA types. The SOFA score and in-hospital mortality were lower for CC1- MRSA-IV, with 2.3 ± 0.6 and 12.5%, respectively than those of other major MRSA types; however, there was no statistical difference.

**Table 2.**
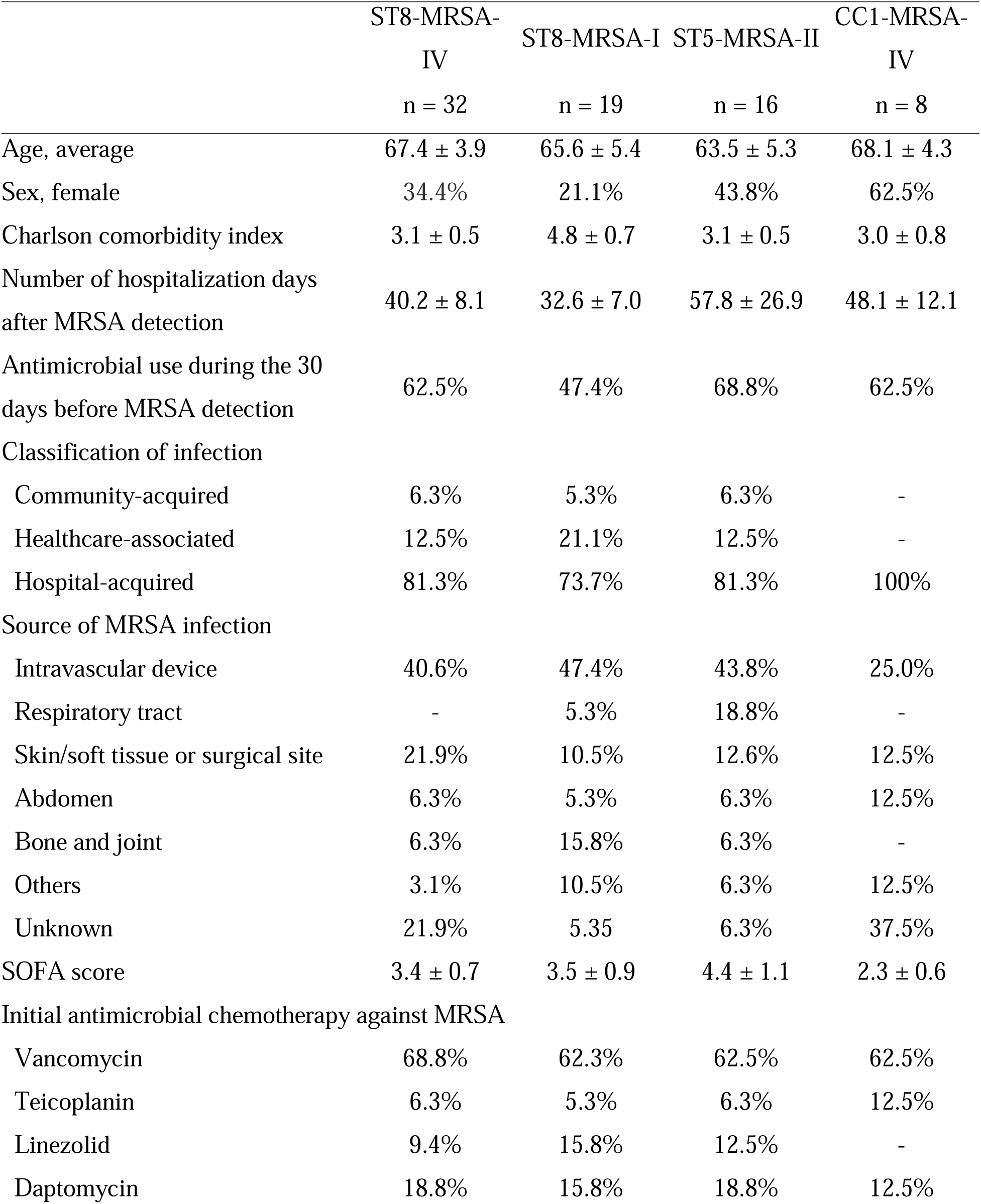
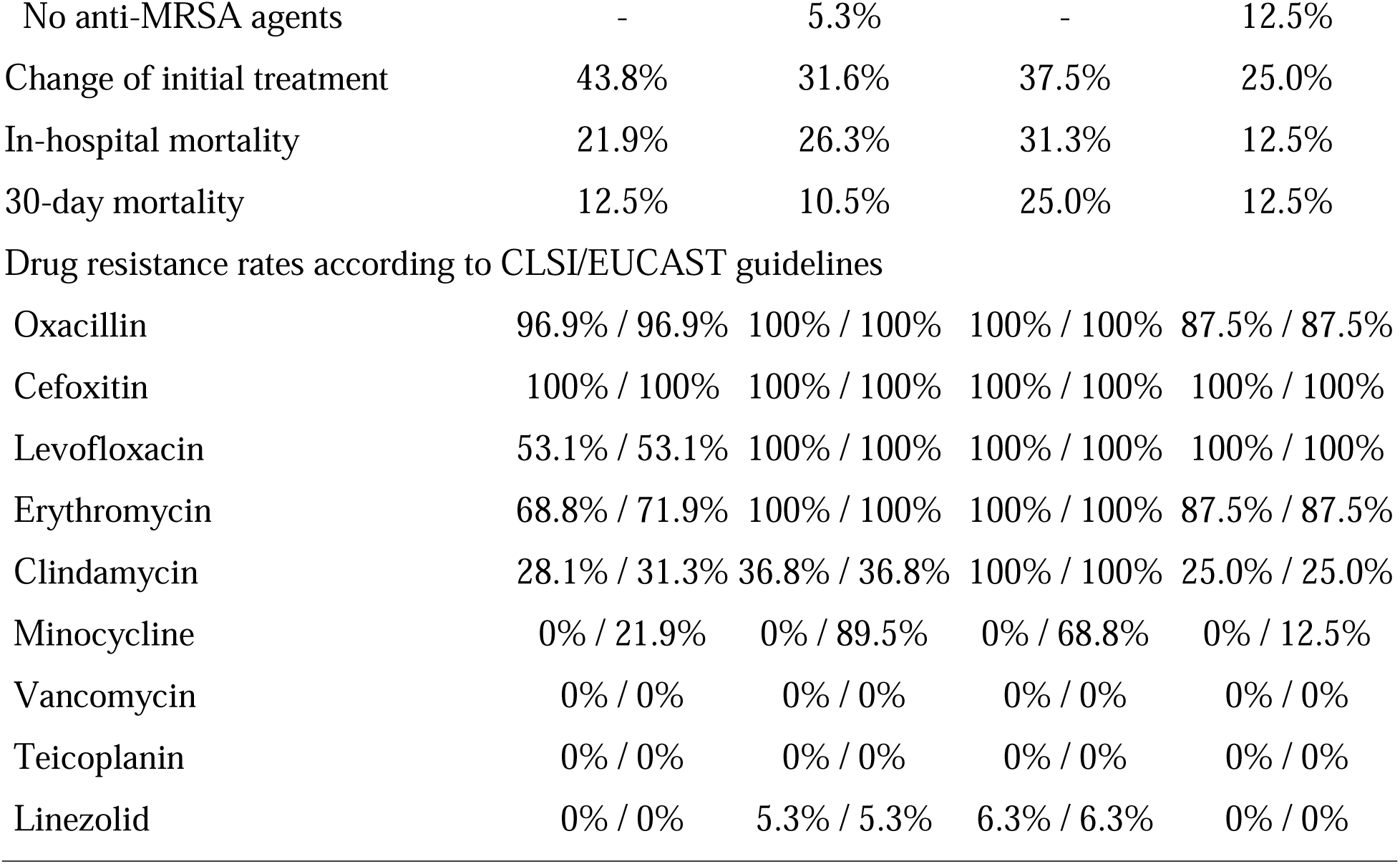
Patient characteristics and drug-resistance rates according to the major MRSA types.

|  | ST8-MRSA-IV<br>n = 32 | ST8-MRSA-I<br>n = 19 | ST5-MRSA-II<br>n = 16 | CC1-MRSA-IV<br>n = 8 |
| --- | --- | --- | --- | --- |
| Age, average | 67.4 ± 3.9 | 65.6 ± 5.4 | 63.5 ± 5.3 | 68.1 ± 4.3 |
| Sex, female | 34.4% | 21.1% | 43.8% | 62.5% |
| Charlson comorbidity index | 3.1 ± 0.5 | 4.8 ± 0.7 | 3.1 ± 0.5 | 3.0 ± 0.8 |
| Number of hospitalization days after MRSA detection | 40.2 ± 8.1 | 32.6 ± 7.0 | 57.8 ± 26.9 | 48.1 ± 12.1 |
| Antimicrobial use during the 30 days before MRSA detection | 62.5% | 47.4% | 68.8% | 62.5% |
| Classification of infection |  |  |  |  |
| Community-acquired | 6.3% | 5.3% | 6.3% | - |
| Healthcare-associated | 12.5% | 21.1% | 12.5% | - |
| Hospital-acquired | 81.3% | 73.7% | 81.3% | 100% |
| Source of MRSA infection |  |  |  |  |
| Intravascular device | 40.6% | 47.4% | 43.8% | 25.0% |
| Respiratory tract | - | 5.3% | 18.8% | - |
| Skin/soft tissue or surgical site | 21.9% | 10.5% | 12.6% | 12.5% |
| Abdomen | 6.3% | 5.3% | 6.3% | 12.5% |
| Bone and joint | 6.3% | 15.8% | 6.3% | - |
| Others | 3.1% | 10.5% | 6.3% | 12.5% |
| Unknown | 21.9% | 5.35 | 6.3% | 37.5% |
| SOFA score | 3.4 ± 0.7 | 3.5 ± 0.9 | 4.4 ± 1.1 | 2.3 ± 0.6 |
| Initial antimicrobial chemotherapy against MRSA |  |  |  |  |
| Vancomycin | 68.8% | 62.3% | 62.5% | 62.5% |
| Teicoplanin | 6.3% | 5.3% | 6.3% | 12.5% |
| Linezolid | 9.4% | 15.8% | 12.5% | - |
| Daptomycin | 18.8% | 15.8% | 18.8% | 12.5% |
| No anti-MRSA agents | - | 5.3% | - | 12.5% |
| Change of initial treatment | 43.8% | 31.6% | 37.5% | 25.0% |
| In-hospital mortality | 21.9% | 26.3% | 31.3% | 12.5% |
| 30-day mortality | 12.5% | 10.5% | 25.0% | 12.5% |
| Drug resistance rates according to CLSI/EUCAST guidelines |  |  |  |  |
| Oxacillin | 96.9% / 96.9% | 100% / 100% | 100% / 100% | 87.5% / 87.5% |
| Cefoxitin | 100% / 100% | 100% / 100% | 100% / 100% | 100% / 100% |
| Levofloxacin | 53.1% / 53.1% | 100% / 100% | 100% / 100% | 100% / 100% |
| Erythromycin | 68.8% / 71.9% | 100% / 100% | 100% / 100% | 87.5% / 87.5% |
| Clindamycin | 28.1% / 31.3% | 36.8% / 36.8% | 100% / 100% | 25.0% / 25.0% |
| Minocycline | 0% / 21.9% | 0% / 89.5% | 0% / 68.8% | 0% / 12.5% |
| Vancomycin | 0% / 0% | 0% / 0% | 0% / 0% | 0% / 0% |
| Teicoplanin | 0% / 0% | 0% / 0% | 0% / 0% | 0% / 0% |
| Linezolid | 0% / 0% | 5.3% / 5.3% | 6.3% / 6.3% | 0% / 0% |

The drug-resistance rates of the major MRSA types to different antimicrobial agents are shown in Table 2 and Supplementary Table 3. The levofloxacin resistance rate was significantly lower in ST8-MRSA-IV than in all the other major MRSA types (*q* = 0.002 vs. ST8-MRSA-I and ST5-MRSA-II and *q* = 0.032 vs. CC1-MRSA-IV). The erythromycin resistance rate was also significantly lower in ST8-MRSA-IV than in ST8-MRSA-I. In contrast, the clindamycin resistance rate of ST5-MRSA-II was significantly higher than that of all other major MRSA types (*q* < 0.001 for all comparisons). CLSI and EUCAST guidelines differ in their protocols for determining minocycline resistance rates. All strains were sensitive to minocycline per the CLSI guidelines; however, when tested according to the EUCAST guidelines, a difference in minocycline resistance rates was observed among the major MRSA types. The minocycline resistance rate was significantly higher in ST8-MRSA-I and ST5-MRSA-II than in ST8-MRSA-IV and CC1-MRSA-IV. There were also differences in the MICs of beta-lactams, for which the CLSI or EUCAST did not set breakpoints for MRSA. The MICs of cefoxitin, cefazolin, imipenem, and meropenem were lower for ST8-MRSA-IV and CC1-MRSA-IV than for ST8- MRSA-I and ST5-MRSA-II (Fig. 3A). Especially, the MICs of imipenem against most strains belonging to the ST8-MRSA-IV and CC1-MRSA-IV were less than 0.5, in contrast to those of the ST8-MRSA-I and ST5-MRSA-II, which were higher than 16 (Fig. 3A).

**Fig. 3.**
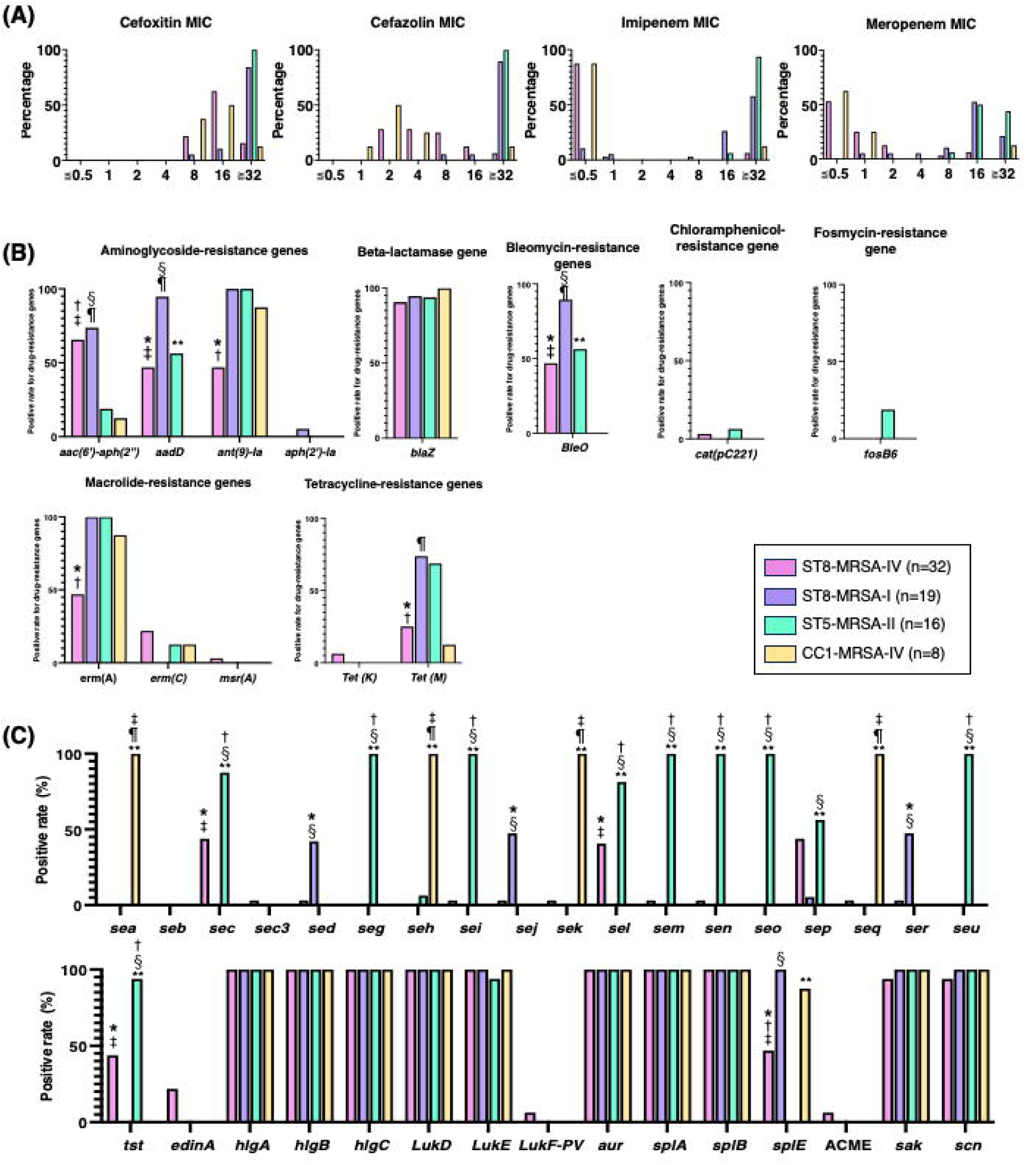
Microbiological characteristics of the major MRSA types. Minimum inhibitory concentration (A) was measured by broth microdilution testing using Dry Plate Eiken (Eiken, Tokyo, Japan) according to the manufacturer’s instructions. Drug-resistance (B) and virulence (C) genes were detected after whole-genome sequencing using ResFinder and virulence finder, respectively. Statistically significant differences were observed in *ST8- MRSA-IV versus ST8-MRSA-I, ^†^ST8-MRSA-IV versus ST5-MRSA-II, ^‡^ST8-MRSA-IV versus CC1-MRSA-IV, ^§^ST8-MRSA-I versus ST5-MRSA-II, ^¶^ ST8-MRSA-I versus CC1-MRSA-IV, and **ST5-MRSA-II versus CC1-MRSA-IV.

### Molecular characteristics of the strains in the major MRSA types

Drug resistance and virulence genes differed among the major MRSA types. Although ST8- MRSA-IV and ST8-MRSA-I belonged to the same sequence type, the positivity rates for drug-resistance genes, such as *aadD, ant(9)-Ia, erm(A), tet(M),* and *bleO,* in ST8-MRSA-I were significantly higher than those in ST8-MRSA-IV (Fig. 3B). ST8-MRSA-I showed a similar trend with respect to its drug-resistance genes as that demonstrated by ST5-MRSA-II, except for *aac(6*′*)-aph(2*′′). However, the positivity rates for the virulence genes differed between ST8- MRSA-I and ST5-MRSA-II. The positivity rates for *sec*, *seg*, *sei*, *sel*, *sem*, *seo*, *sep*, *seu*, and *tst* were significantly higher in ST5-MRSA-II than in ST8-MRSA-I. In contrast, the positivity rate for *splE* was significantly lower in ST5-MRSA-II than in ST8-MRSA-I (Fig. 3C). CC1-MRSA-IV has characteristic virulence gene features. It harbors virulence genes such as *sea*, *seh*, *sek*, and *seq,* which are absent in other major MRSA types.

In ST8-MRSA-IV, the positivity rates for drug-resistance and virulence genes, such as *aac(6*^′^*)- aph(2*^′′^), *aadD*, *ant(9)-Ia*, *erm(A)*, *bleO*, *sec*, *sel*, *sep*, *tst*, and *splE*, were around 50% (Fig. 3B and C). Phylogenetic analysis based on core genome MLST revealed that ST8-MRSA-IV was divided into several clusters (Fig. 4). Based on the results of the virulence genes *sec* and *tst* and the *spa* type,^5^ 14 and 4 strains were classified as CA-MRSA/J and t5071-ST8-MRSA-IV, respectively. There were significant differences between CA-MRSA/J and t5071-ST8-MRSA-IV gene expression (Supplementary Table 4). The positive rates for *aac(6*^′^*)-aph(2*^′′^*), aadD, bleO, sec, sel,* and *tst* were significantly higher in CA-MRSA/J than in t5071-ST8-MRSA-IV (*p* < 0.05), and those for *ant(9)-Ia, erm(A), splE,* and *sep* were significantly lower in CA-MRSA/J than in t5071-ST8-MRSA-IV (*p* < 0.05). Based on antimicrobial susceptibility testing, CA-MRSA/J was more sensitive to levofloxacin, erythromycin, clindamycin, and minocycline than was t5071-ST8-MRSA-IV (*p* < 0.05) (Supplementary Table 4).

**Fig. 4.**
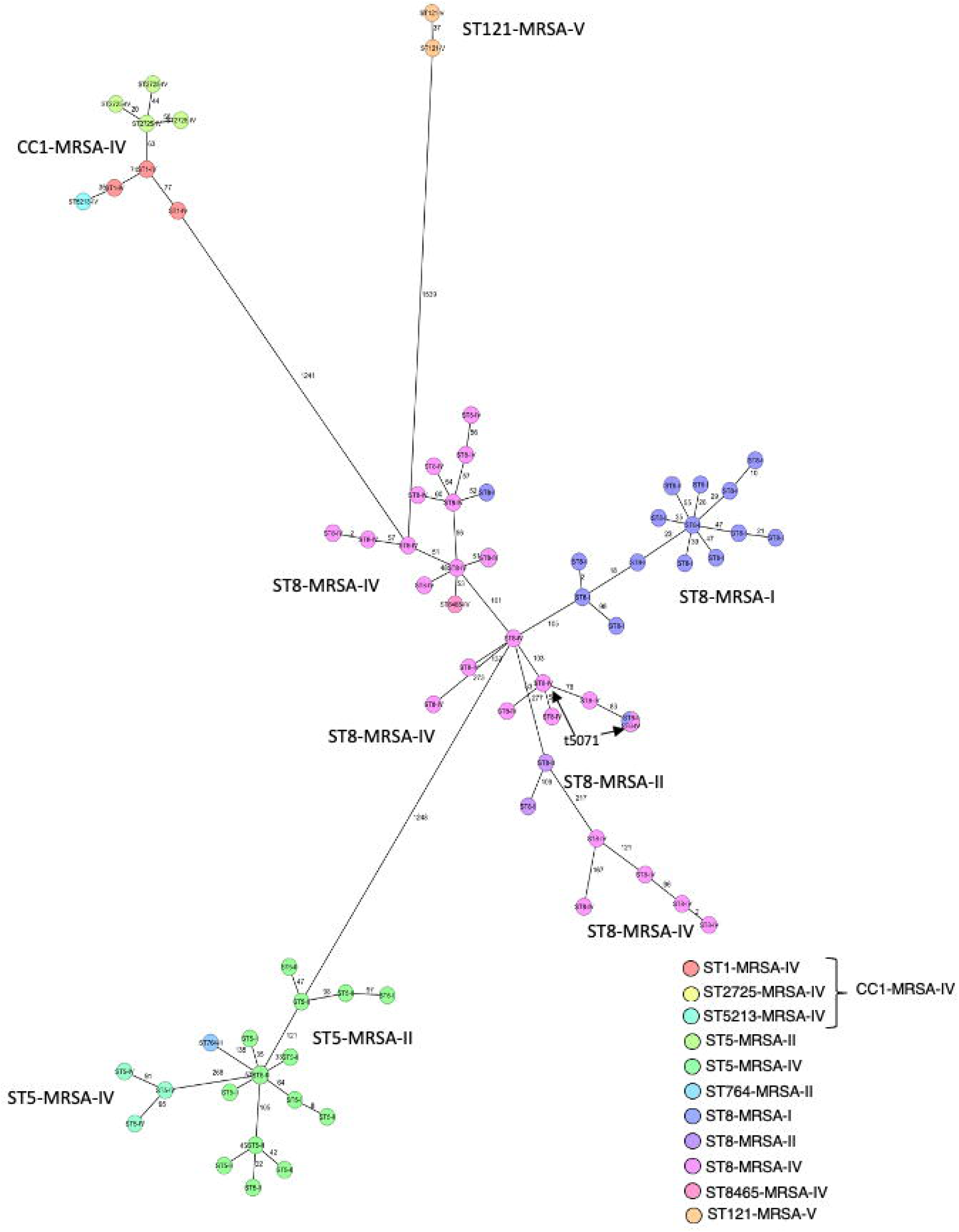
Phylogenetic analysis based on core genome MLST. MLST (cgMLST) was performed using Ridom SeqSphere+ v.9.0.10 (Ridom GmbH, Münster, Germany). A minimum spanning tree (MST) was created based on MLST, cgMLST, and S. aureus Accessory using Ridom SeqSphere+ (ver.9). Samples with more than 10% missing values of the items for distance calculation were excluded from the MST.

### Relationship between the strains detected in Nagasaki and those circulating in Japan

Phylogenetic tree analysis based on MLST, cgMLST, and *S. aureus* accessory sequences was performed to investigate the relationship between the strains detected in this study and those from the previous nationwide surveillance.^5^ Based on the phylogenetic tree analysis, the strains were divided into three major clusters, ST8-MRSA-IV and ST8-MRSA-I, CC1-MRSA-IV, and ST5-MRSA-II (Supplementary Fig. 2). In the ST8-MRSA-IV and ST8-MRSA-I clusters (Fig. 5A), most ST8-MRSA-I strains detected in Nagasaki and the nationwide surveillance were classified into subcluster 3, whereas ST8-MRSA-IV was divided into ten subgroups. However, in the ST8-MRSA-IV group, all CA-MRSA/J strains detected in Nagasaki and nationwide were classified into subcluster 1. All t5071-ST8-MRSA-IV strains detected in Nagasaki and nationwide were classified into subcluster 5. The ST5-MRSA-II strains showed the same trend as that of the ST8-MRSA-IV strains. The ST5-MRSA-II strains detected in Nagasaki were divided into five subclusters (Fig. 5C). In contrast to ST8-MRSA-IV and ST5-MRSA-II, most CC1- MRSA-IV strains detected in Nagasaki and nationwide surveillance were classified into the same sub-cluster 12 (Fig. 5B).

**Fig. 5.**
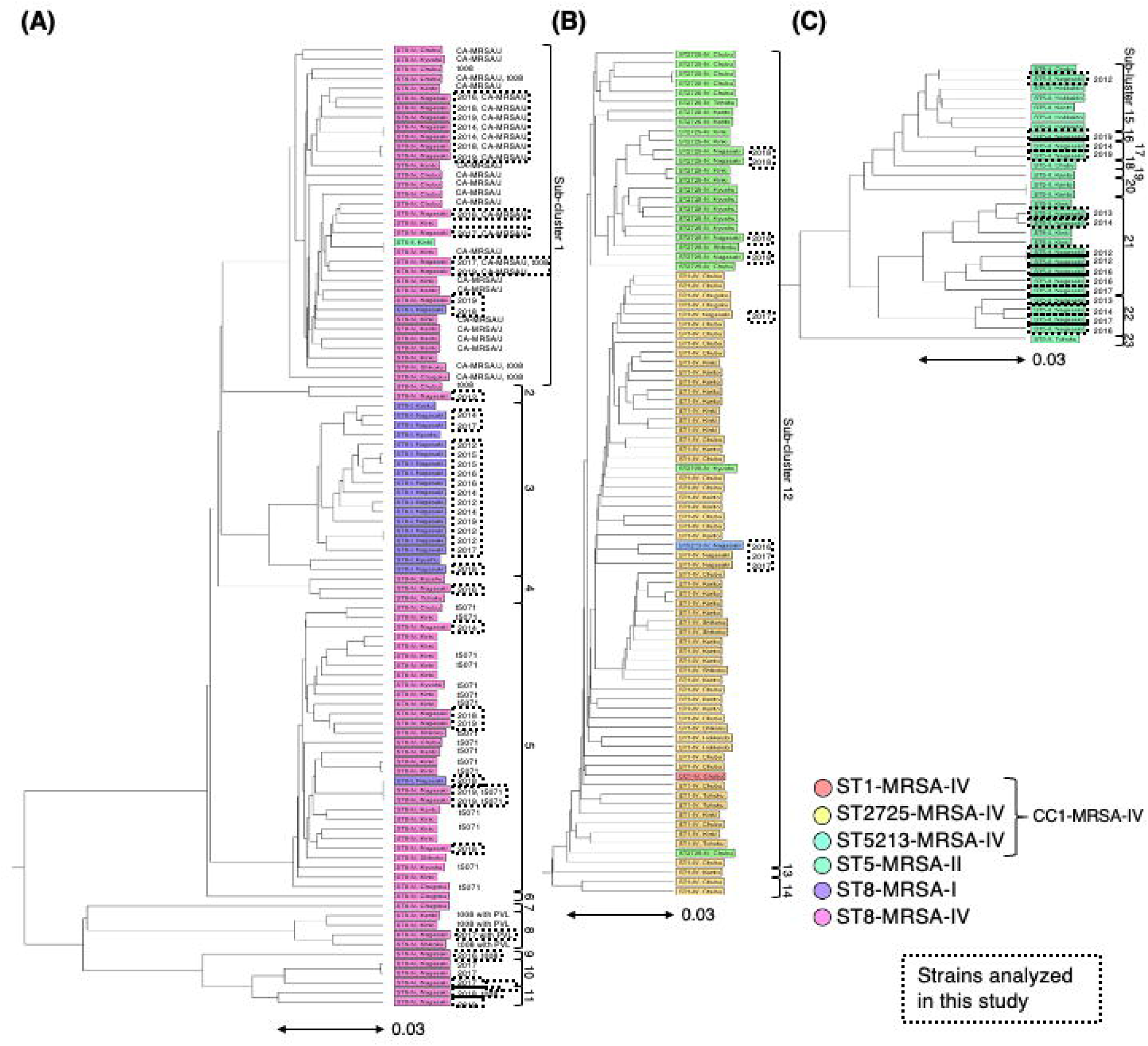
Phylogenetic analysis of MRSA strains detected in Nagasaki and those circulating in Japan. Phylogenetic tree analysis was performed based on MLST, core genome MLST (cgMLST), and *S. aureus* accessory sequences to investigate the relationship between the strains detected in this study and those from the previous nationwide surveillance. Based on the phylogenetic tree analysis of all strains (Supplementary Fig. 2), the strains were divided into three major clusters: ST8-MRSA-IV and ST8-MRSA-I (**A**), CC1-MRSA-IV (**B**), and ST5-MRSA-II (**C**). cgMLST was performed using the Ridom SeqSphere+ v.9.0.10 (Ridom GmbH, Münster, Germany). A UPGMA tree was created based on MLST, cgMLST, and *S. aureus* accessory genes. Samples with more than 10% missing values among the data used for distance calculation were excluded. During UPGMA of the strains detected during the previous nationwide surveillance, the strains detected at Nagasaki University Hospital were excluded to avoid duplication.

## Discussion

This study revealed that SCC*mec* type IV has quickly become the predominant SCC*mec* type and replaced SCC*mec* type II since the mid-2010s in patients with BSIs. In the United States, SCC*mec* type IV has increased prevalence and replaced SCC*mec* type II in BSIs since the late 2000s.^15^ Based on the results from the two nationwide surveillance conducted in Japan, the prevalence of SCC*mec* type IV increased from 19.9% in 2011 to 77.4% in 2019, whereas that of SCC*mec* type II decreased to 7.8% in 2019 from 75.6% in 2011.^6,16^ In addition, according to a previous Japanese study conducted in the Kanto region, the prevalence of SCC*mec* type IV was higher than that of SCC*mec* type II in 2016,^17^ which is similar to the results of this study. These results indicate that the shift from SCC*mec* type II to IV occurred almost simultaneously throughout Japan in the mid-2010s. Although SCC*mec* type IV is the predominant SCC*mec* type in both the United States and Japan, its molecular characteristics differ. The USA300-like clone (t008-ST8-MRSA-IV) was the most common MRSA type in the United States.^15^ The USA300- like clone carries Panton-Valentine leucocidin (PVL) and arginine catabolic mobile element (ACME) genes, contributing to its superior virulence and adaptability compared to ST5-MRSA-II, making it the predominant MRSA clone in the United States.^18^ However, the percentage of the USA300-like clone was only 3.3% in Japan in 2019,^6^ which is similar to the results of this study. In both the Japanese nationwide study and our study, ST8-MRSA-IV and CC1-MRSA-IV without PVL and ACME replaced ST5-MRSA-II.^6^ These clones have fewer virulence genes and better antimicrobial susceptibility than ST5-MRSA-II. However, a previous study reported that SCCmec type IV strains isolated in Japan have a higher plasma-biofilm formation ability than SCCmec type II strains.^17^ In addition, the positive rate of *splE*, a gene associated with the interaction with host proteins during infection,^19^ was significantly higher in ST8-MRSA-IV and CC1-MRSA-IV than in ST5-MRSA-II in our study. Thus, the increase in SCCmec type IV in Japan is likely driven by different mechanisms compared to the one observed in the USA300-like clone. The emergence and spread of SCC*mec* type IV clones are not confined to Japan and the United States. In other parts of Asia, distinct SCCmec type IV MRSA clones have also been identified. For instance, in South Korea, by 2018, ST72-MRSA-IV replaced the previously dominant clone ST5-MRSA-II and ST239-MRSA-III.^20^ Similarly, in southern China, ST59- MRSA-IV was the most common MRSA clone in pediatrics.^21^ These findings suggest that the evolution and dissemination of SCC*mec* type IV MRSA clones vary significantly across the country, driven by local epidemiological and ecological factors.

We also investigated the changes in patient characteristics. Although patient backgrounds did not change between 2003 and 2019, SOFA scores and in-hospital mortality improved significantly. In addition, the source of MRSA infection has changed. The percentage of patients with intravascular device-related BSI was much higher during 2016–2019 than during 2003–2007, whereas that of patients with respiratory tract infections was much lower during 2016– 2019 than during 2003–2007. We also compared the patient characteristics among the major MRSA types. The SOFA score and in-hospital mortality in patients with the ST5-MRSA-II were higher than those with ST8-MRSA-IV, ST8-MRSA-I, or CC1-MRSA-IV types. ST5-MRSA-II exhibited a high prevalence of several toxins, such as *sec, seg, sei, sem, sen, seo*, *seu,* and *tst*. In addition, the percentage of patients with respiratory tract infections was higher in ST5-MRSA-II than in ST8-MRSA-IV, ST8-MRSA-I, and CC1-MRSA-IV. In the *S. aureus* BSIs, pneumonia was reported as an independent factor associated with death.^22^ These factors likely contributed to the higher SOFA score and in-hospital mortality in patients with the ST5-MRSA-II compared to other MRSA types. In contrast, the SOFA score and in-hospital mortality in patients with CC1- MRSA-IV were lower than those with other MRSA types. Because the prevalence of ST5- MRSA-II decreased and those of ST8-MRSA-IV and CC1-MRSA-IV increased from 2003 to 2019, and there was no change in patient background, the changes in MRSA types may have reduced the severity and in-hospital mortality.

CA-MRSA/J, t5071-ST8-MRSA-IV, and CC1-MRSA-IV, the major MRSA types in Japan in 2019,^6,^ were also detected in this study and classified into distinct clusters in the phylogenetic analysis. Furthermore, the strains detected in this study ^6^were classified into the same subcluster as those detected in the Japanese nationwide surveillance.^6^ In addition, The drug susceptibility characteristic trends in this study were similar to those in the national surveillance^6^ for most cases. The following differences among the three major types were observed in both studies. CA-MRSA/J was sensitive to levofloxacin; CA-MRSA/J and CC1-MRSA-IV were sensitive to clindamycin, while t5071-ST8-MRSA-IV was resistant; for minocycline, all types were sensitive according to CLSI criteria, whereas t5071-ST8-MRSA-IV had a very high rate of resistance according to the EUCAST criteria. The positivity rates for drug-resistance genes showed the same trends in both studies. Almost all CC1-MRSA-IV and t5071-ST8-MRSA-IV harbored *ant(9)-Ia* and *erm* (A). In contrast, the positivity rates for *aac(6*^′^*)-aph(2*^′′^) and *aadD* were much higher in CA-MRSA/J than in other types. The positivity rates for virulence genes showed the same trend in both studies. The virulence genes *sea, seh, sek,* and *seq* were harbored by the CC1- MRSA-IV but not the other two types. Similarly, CA-MRSA/J, but not the other two types, harbored *sec, sel,* and *tst*. In contrast, CA-MRSA/J did not harbor *splE*. These results suggest that CA-MRSA/J, t5071-ST8-MRSA-IV, and CC1-MRSA-IV have spread across the nation rapidly with the same molecular background and characteristics.

However, there were several differences in the percentage of CA-MRSA/J, t-571-ST8-MRSA-IV, and CC1-MRSA-IV between this study and Japanese nationwide surveillance.^6^ In this study, although CA-MRSA/J was the predominant strain of ST8-MRSA-IV, t5071-ST8-MRSA-IV was not frequently detected. CA-MRSA/J harbors the superantigenic toxin-encoding *S. aureus* pathogenicity island (SaPI), which includes *sec, tst*, and *sel* genes. This clone has been widely reported in Japan since the 2000s, primarily linked to skin infections.^23^ In contrast, t5071-ST8- MRSA-IV was first identified as a spreading strain in Japan in the previous nationwide surveillance ^6^ and was only detected only after 2016 in this study. A similar phenomenon was also observed in CC1-MRSA-IV, detected only after 2016. ^66^Although there were regional differences in the ratios of CC1-MRSA-IV and ST8-MRSA-IV in Japan, the percentage of ST8- MRSA-IV was much higher than that of CC1-MRSA-IV in western Japan, including the Kyushu region, where Nagasaki belongs.^6^ However, in a single-center study conducted in the Kyushu region during 2018–2019, the percentage of CC1-MRSA-IV was 14.2% in blood cultures, whereas it was 59.0% and 77.3% in sputum and skin and soft tissues, respectively.^24^ In addition, a previous study conducted in Hokkaido reported a rapidly increasing percentage of CC1- MRSA-IV in BSIs in 2019.^25^ In addition, a study on MRSA bloodstream infections in the Kyushu region reported a notable increase in CC1-MRSA-IV after 2016, with a marked rise especially after 2019.^26^ Both t5071-ST8-MRSA-IV and CC1-MRSA-IV had a high positive rate for splE gene^,^ distinguishing them from CA-MRSA/J, which lacked splE similar to ST5-MRSA-II.^6^ Since both t5071-ST8-MRSA-IV and CC1-MRSA-IV were detected only after 2016 in this study, and the strains within each clone were highly similar in MST analysis, it is possible that we only observed the early stages of these clones’ circulation in our region.

The present study also revealed that a different epidemic strain was prevalent in our hospital compared to those identified in the nationwide surveillance. In our hospital, the prevalence of SCC*mec* type I increased from 2003 to 2015 and became the predominant MRSA type during 2012-2015. Unlike the ST8-MRSA-IV, ST8-MRSA-I showed a similar pattern to ST5-MRSA-II in drug resistance. The difference from ST5-MRSA-II was that ST8-MRSA-I was more sensitive to clindamycin and exhibited higher positivity rates for *aac(6*^′^*)-aph(2*^′′^) and *aadD.* In contrast, their virulence genes were completely different, with ST8-MRSA-I having markedly fewer virulence factors than ST5-MRSA-II. In virulence genes, ST8-MRSA-I showed a trend similar to that of t5071-ST8-MRSA-IV; however, *sed, sej,* and *ser* were detected only in ST8-MRSA-I. In the nationwide surveillance study, four ST8-MRSA-I strains were detected in the Kyushu region.^6^ In a previous single-center study in the Kyushu region, ST8-MRSA-I was the second most frequently detected type during 2018–2019.^24^ Several studies in the Kyushu region detected SCC*mec* type I strains at certain rates: 8.9% during 2005-2011,^27^ 14.5% during 2014-2015,^28^ and 9.3% during 2019-2020. ^29^ These results indicated that ST8-MRSA-I spread independently in the Kyushu region. In this study, the prevalence of ST8-MRSA-I decreased from 37.0% during 2012–2015 to 15.5% during 2016–2019. A similar trend was observed in a study on MRSA BSI in the Kyushu region, where the proportion of SCCmec type I decreased from 32% in 2013–2015 to 10% in 2016–2018 and further dropped to 6% in 2019–2021.^26^ As previously mentioned, ST8- MRSA-I exhibited high susceptibility to clindamycin; however, the Antimicrobial Use Density (AUD) for clindamycin at our hospital remained relatively stable, at 0.32 in 2012 and 0.29 in 2019, indicating no significant increase. While ST8-MRSA-I frequently harbors the resistance genes *aac(6’)-aph(2’’)* and *aadD*, these genes are rarely found in CC1-MRSA-IV. At our hospital, the AUD for aminoglycosides decreased significantly from 0.45 in 2012 to 0.14 in 2019, suggesting that reduced aminoglycoside use may have contributed to the replacement of the endemic ST8-MRSA-I by SCCmec type IV strains, such as CC1-MRSA-IV, in the Kyushu region.

This study has several limitations. First, because this was a single-center study, it is unclear whether the differences in patient characteristics for each MRSA type can be generalized. Second, to reduce the likelihood of contamination, we included only cases that were positive for at least two sets of blood cultures. This inclusion criterion is the same as that used in the nationwide surveillance^5^ but different from our previous studies.^4,5^ Under the previous criteria, in-hospital mortality during 2012-2015 would have been lower than the current study (24.3% vs 39.8%), while it was similar for 2016-2019 (17.0% vs 15.5%). Thus, we believe the observed mortality reduction is not due to the change in the inclusion criteria. In contrast, the previous inclusion criteria would have yielded 70 strains for 2012-2015 and 94 strains for 2016-2019, indicating a significant reduction in the number of strains analyzed in 2012-2015 due to the criteria change. However, since the decision to collect one or two sets of blood cultures was left to the clinicians’ discretion, we believe this decision is unlikely to have influenced the distribution of MRSA clones. Third, in this study, we aimed to investigate the changes at our institution up to the year of the nationwide surveillance conducted in Japan, which is why we limited our analysis to strains collected until 2019. Further changes in MRSA clones may have occurred since 2020, influenced by the COVID-19 pandemic and other factors. Therefore, ongoing surveillance and further investigation will be necessary to monitor these potential shifts. Fourth, there were several strains for which the *spa* type was not determined or excluded from the phylogenetic analysis: 22 of 85 strains (25.9%) and 10 of 75 strains (13.3%), respectively. This is a limitation that also holds for nationwide surveillance.^5^ Lastly, neither this study nor the previous nationwide surveillance included evolutionary phylogenetic analysis to estimate when the major MRSA clones emerged. Future studies incorporating more extensive genomic data from across Japan are needed to address this limitation.

In conclusion, this study demonstrated that the major MRSA types in BSIs changed over time and were associated with reduced disease severity and in-hospital mortality. In addition, changes in the major MRSA types were directly influenced by changes in the circulating strains nationally and regionally.

## Data availability

Raw data were generated at the Nagasaki University Hospital. The derived data supporting the findings of this study are presented in the paper and supplementary tables and figures. The WGS data reported in this study are available in the NCBI Sequence Read Archive (https://www.ncbi.nlm.nih.gov/sra) under the accession number PRJNA1065106. WGS data from the previous nationwide surveillance^5^ are available in the DDBJ Sequence Read Archive (https://www.ddbj.nig.ac.jp/dra/index-e.html) under the accession number DRA013058.

## Conflicts of interest

The authors have no conflicts to declare.

## Funding

This work was supported by the Japanese Association for Infectious Diseases (The 1^st^ Grant for Clinical Research Promotion, 2018) and the Japanese Antibiotics Research Association (The 23^rd^ Encouragement Award, 2021). The funders had no role in the data analysis or the decision to publish.

## Author contributions

N. K. and K. Y. designed the experiments. N. K., M. I., G. Y., and D. S. acquired the data. N.K., K.O., F. M. K., K.K., H.H., K.I., H.M., and K.Y. analyzed the data. N.K. wrote the manuscript. All the authors have revised and approved the manuscript for publication.

## Acknowledgments

We thank Mr. Shuji Miyazaki (Department of Laboratory Medicine, Nagasaki University of Hospital) for supporting the analysis of MRSA strains. We would like to thank Editage (www.editage.jp) for editing the English language.

## Notes

### Competing Interest Statement

The authors have declared no competing interest.

### Funding Statement

This work was supported by the Japanese Association for Infectious Diseases (The 1st Grant for Clinical Research Promotion, 2018) and the Japanese Antibiotics Research Association (The 23rd Encouragement Award, 2021). The funders had no role in the data analysis or the decision to publish.

### Summary of Updates

We revised the aim of the study and discussion section to clarify the important point of this manuscript. Figures and supplementary Figures were also revised.

