## Supplementary material for "Long-Term Impact of Molecular Epidemiology Shifts of Methicillin-resistant *Staphylococcus aureus* on Severity and Mortality of Bloodstream Infection": Supplemantary Figure 1

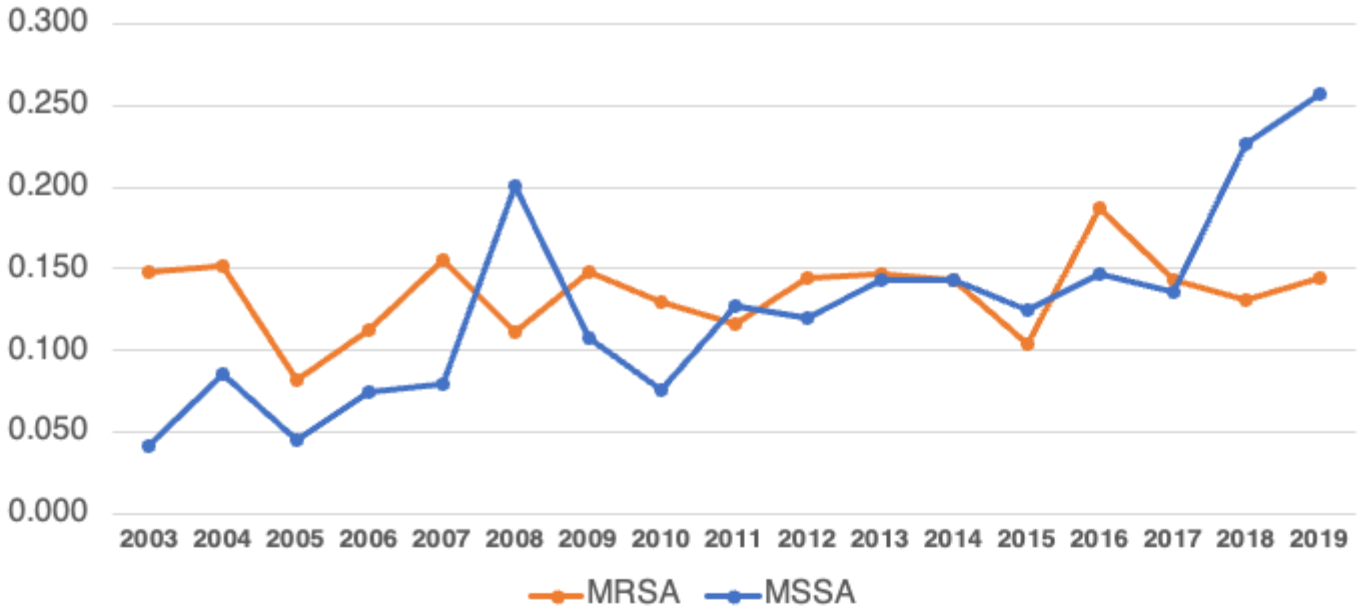


**Supplementary Fig. 1. Number of isolates detected in blood cultures**

The numbers are expressed per 1,000 patient days. Duplicates were excluded when isolates were obtained from the same patient on the same day.
