## Supplementary material for "Long-Term Impact of Molecular Epidemiology Shifts of Methicillin-resistant *Staphylococcus aureus* on Severity and Mortality of Bloodstream Infection": Supplemantary Figure 2

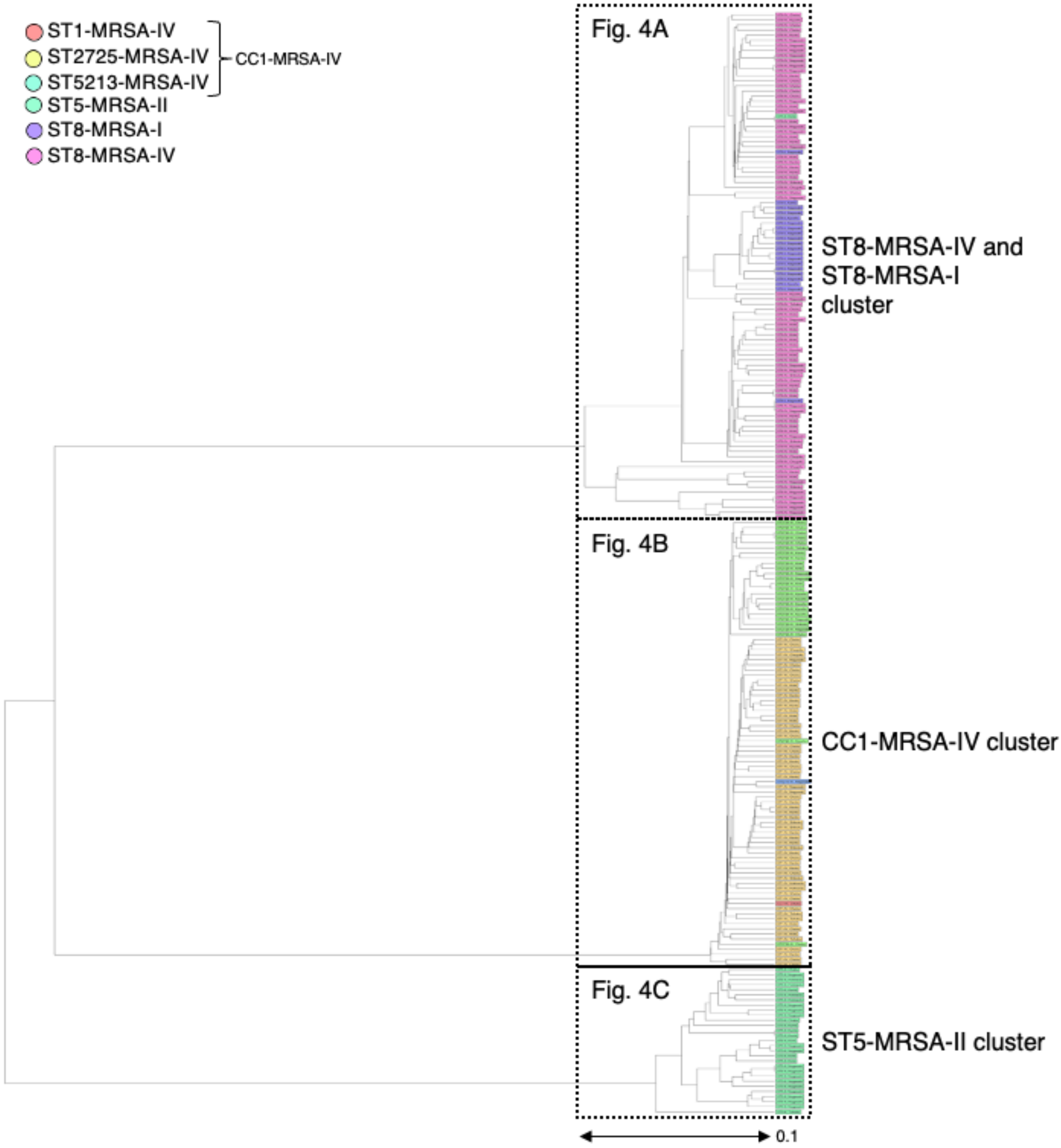


**Supplementary Fig. 2. Phylogenetic analysis of MRSA strains collected in this study and nationwide surveillance based on core genome MLST**

MLST (cgMLST) was performed using Ridom SeqSphere+ v.9.0.10 (Ridom GmbH, Münster, Germany). UPGMA tree was created based on MLST, cgMLST, and S. aureus accessory genes. UPGMA tree was created based on MLST, cgMLST, and S. aureus accessory genes. Samples with more than 10% missing values of the items for distance calculation were excluded. In the UPGMA analysis using the strains from the previous nationwide surveillance, the strains detected in Nagasaki University Hospital were excluded from the strains to avoid duplication.
