## Supplementary material for "Long-Term Impact of Molecular Epidemiology Shifts of Methicillin-resistant *Staphylococcus aureus* on Severity and Mortality of Bloodstream Infection": Table S1

Supplementary Table 1.Changes in SCC*mec* types and patient characteristics from 2003 to 2019

| | 2003-2007 (n=83) | 2008-2011 (n=83) | 2012-2015 (n=27) | 2016-2019 (n=58) | 2003-2007 vs 2008-2011 | | 2003-2007 vs 2012-2015 | | 2003-2007 vs 2016-2019 | | 2008-2011 vs 2012-2015 | | 2008-2011 vs 2016-2019 | | 2012-2015 vs 2016-2019 | |
|---|---|---|---|---|---|---|---|---|---|---|---|---|---|---|---|---|
| | n (%) | n (%) | n (%) | n (%) | *P* value | *Q* value | *P* value | *Q* value | *P* value | *Q* value | *P* value | *Q* value | *P* value | *Q* value | *P* value | *Q* value |
| SCC*mec* types | | | | | | | | | | | | | | | | |
| SCC*mec* type I | 2 (2.6%) | 18 (23.1%) | 10 (37.0%) | 9 (15.5%) | <0.001 | <0.001 | <0.001 | <0.001 | 0.009 | 0.019 | n.s. | | n.s. | | 0.048 | 0.072 |
| SCC*mec* type II | 61 (79.2%) | 35 (44.9%) | 10 (37.0%) | 9 (15.5%) | <0.001 | <0.001 | <0.001 | <0.001 | <0.001 | <0.001 | n.s. | | 0.000 | 0.001 | 0.048 | 0.057 |
| SCC*mec* type III | - - | 1 (1.3%) | - - | - - | n.s. | | n.s. | | n.s. | | n.s. | | n.s. | | n.s. | |
| SCC*mec* type IV | 14 (18.2%) | 24 (30.8%) | 7 (25.9%) | 38 (65.5%) | 0.092 | 0.139 | n.s. | | <0.001 | <0.001 | n.s. | | <0.001 | <0.001 | 0.001 | 0.002 |
| SCC*mec* type IV | - - | - - | - - | 2 (3.4%) | n.s. | | n.s. | | 0.183 | n.s. | n.s. | | 0.180 | n.s. | n.s. | |
| Patient characteristics | | | | | | | | | | | | | | | | |
| Age, average | 63.7 ± 1.9 | 67.3 ± 1.6 | 69.1 ± 3.2 | 63.4 ± 3.2 | n.s. | | n.s. | | n.s. | | n.s. | | n.s. | | n.s. | |
| Sex, female | 39 (47.0%) | 25 (30.1%) | 7 (25.9%) | 21 (36.2%) | 0.038 | n.s. | n.s. | | n.s. | | n.s. | | n.s. | | n.s. | |
| Charlson comorbidity index | 3.0 ± 0.2 | 3.1 ± 0.2 | 4.0 ± 0.5 | 3.2 ± 0.3 | n.s. | | n.s. | | n.s. | | n.s. | | n.s. | | n.s. | |
| Number of days of hospitalization after MRSA detection | No data | 41.9 ± 6.3 | 38.5 ± 6.3 | 36.1 ± 7.9 | - | - | - | - | - | - | n.s. | - | n.s. | - | n.s. | - |
| Antimicrobial use during the 30 days before MRSA detection | No data | 63 (75.9%) | 16 (59.3%) | 35 (60.3%) | - | - | - | - | - | - | 0.138 | n.s. | 0.063 | 0.189 | n.s. | |
| Classification of infection | | | | | | | | | | | | | | | | |
| Community-acquired | No data | 3 (3.6%) | 0 (0.0%) | 5 (8.6%) | - | - | - | - | - | - | n.s. | | n.s. | | 0.173 | n.s. |
| Healthcare-associated | No data | 3 (3.6%) | 6 (22.2%) | 8 (13.8%) | - | - | - | - | - | - | 0.007 | 0.020 | 0.051 | 0.076 | n.s. | |
| Hospital-acquired | No data | 72 (86.7%) | 21 (77.8%) | 45 (77.6%) | - | - | - | - | - | - | n.s. | | 0.176 | n.s. | n.s. | |
| Source of MRSA infection | | | | | | | | | | | | | | | | |
| Intravascular device | 15 (18.1%) | 25 (30.1%) | 8 (29.6%) | 27 (46.6%) | 0.102 | n.s. | n.s. | | <0.001 | 0.002 | n.s. | | 0.053 | 0.158 | 0.162 | 0.195 |
| Respiratory tract | 14 (16.9%) | 17 (20.5%) | 3 (11.1%) | 1 (1.7%) | n.s. | | n.s. | | 0.004 | 0.013 | n.s. | | 0.001 | 0.004 | 0.093 | 0.185 |
| Skin/soft tissue or surgical site | 9 (10.8%) | 13 (15.7%) | 7 (25.9%) | 8 (13.8%) | n.s. | | 0.065 | n.s. | n.s. | | n.s. | | n.s. | | n.s. | |
| Abdomen | 6 (7.2%) | 4 (4.8%) | 0 (0.0%) | 5 (8.6%) | n.s. | | n.s. | | n.s. | | n.s. | | n.s. | | n.s. | |
| Bone and joint | 11 (13.3%) | 2 (2.4%) | 3 (11.1%) | 5 (8.6%) | 0.018 | 0.106 | n.s. | | n.s. | | 0.094 | n.s. | 0.124 | n.s. | n.s. | |
| Others | 6 (7.2%) | 5 (6.0%) | 3 (11.1%) | 2 (3.4%) | n.s. | | n.s. | | n.s. | | n.s. | | n.s. | | n.s. | |
| Unknown | 22 (26.5%) | 17 (20.5%) | 3 (11.1%) | 10 (17.2%) | n.s. | | 0.118 | n.s. | n.s. | | n.s. | | n.s. | | n.s. | |
| SOFA score | 5.8 ± 0.5 | 5.8 ± 0.5 | 4.2 ± 0.9 | 3.1 ± 0.4 | n.s. | | n.s. | | 0.004 | | n.s. | | 0.005 | | n.s. | |
| Initial antimicrobial chemotherapy against MRSA | | | | | | | | | | | | | | | | |
| Vancomycin | 24 (28.9%) | 38 (45.8%) | 15 (55.6%) | 40 (69.0%) | 0.037 | 0.055 | 0.020 | 0.039 | <0.001 | <0.001 | n.s. | | 0.010 | 0.029 | n.s. | |
| Teicoplanin | 33 (39.8%) | 11 (13.3%) | 3 (11.1%) | 3 (5.2%) | 0.000 | 0.001 | 0.008 | 0.017 | <0.001 | <0.001 | n.s. | | 0.155 | n.s. | n.s. | |
| Linezolid | 9 (10.8%) | 18 (21.7%) | 7 0.259 | 1 (1.7%) | 0.091 | 0.109 | 0.065 | 0.098 | 0.047 | 0.094 | n.s. | | <0.001 | 0.002 | 0.001 | 0.003 |
| Daptomycin | 0 (0.0%) | 0 (0.0%) | 3 (11.1%) | 12 (20.7%) | n.s | | 0.014 | 0.027 | <0.001 | <0.001 | 0.014 | 0.027 | <0.001 | <0.001 | n.s. | |
| Arbekacin | 3 (3.6%) | 1 (1.2%) | 0 (0.0%) | 0 (0.0%) | n.s. | | n.s. | | n.s. | | n.s. | | n.s. | | n.s. | |
| No anti-MRSA agents | 14 (16.9%) | 16 (19.3%) | 0 (0.0%) | 3 (5.2%) | n.s. | | 0.020 | 0.059 | 0.039 | 0.058 | 0.010 | 0.062 | 0.022 | 0.045 | n.s. | |
| Change of initial treatment | 29 (34.9%) | 25 (30.1%) | 7 (25.9%) | 22 (37.9%) | n.s. | | n.s. | | n.s. | | n.s. | | n.s. | | n.s. | |
| In-hospital mortality | 33 (39.8%) | 21 (25.3%) | 10 (37.0%) | 9 (15.5%) | 0.068 | 0.136 | n.s. | | 0.003 | 0.015 | n.s. | | n.s. | | 0.048 | 0.143 |
| 30-days mortality | No data | 15 (18.1%) | 5 (18.5%) | 6 (10.3%) | - | - | - | - | - | - | n.s. | | n.s. | | n.s. | |

n.s., *P* or *Q values* > 0.2
