## Supplementary material for "Long-Term Impact of Molecular Epidemiology Shifts of Methicillin-resistant *Staphylococcus aureus* on Severity and Mortality of Bloodstream Infection": Table S2

Supplementary Table 2. Combination of ST and SCC*mec* type in 2012-2019

| | All (n=85) | 2012-2015 (n=27) | | 2016-2019 (n=58) | | *P* value 2012-2015 vs 2016-2019 |
|---|---|---|---|---|---|---|
| | n (%) | n | (%) | n | (%) | |
| CC1-MRSA-IV | 8 (9.4%) | - | - | 8 | (13.8%) | 0.051 |
| ST1-MRSA-IV | 3 (3.5%) | - | - | 3 | (5.2%) | n.s. |
| ST2725-MRSA-IV | 4 (4.7%) | - | - | 4 | (6.9%) | n.s. |
| ST5213-MRSA-IV | 1 (1.2%) | - | - | 1 | (1.7%) | n.s. |
| CC5-MRSA-II | 17 (20.0%) | 9 | (33.3%) | 8 | (13.8%) | 0.045 |
| ST5-MRSA-II | 16 (18.8%) | 8 | (29.6%) | 8 | (13.8%) | 0.134 |
| ST764-MRSA-II | 1 (1.2%) | 1 | (3.7%) | - | - | n.s. |
| CC5-MRSA-IV | 3 (3.5%) | 1 | (3.7%) | 2 | (3.4%) | n.s. |
| ST5-MRSA-IV | 3 (3.5%) | 1 | (3.7%) | 2 | (3.4%) | n.s. |
| CC8-MRSA-I | 19 (22.4%) | 10 | (37.0%) | 9 | (15.5%) | 0.048 |
| ST8-MRSA-I | 19 (22.4%) | 10 | (37.0%) | 9 | (15.5%) | 0.048 |
| CC8-MRSA-II | 2 (2.4%) | 1 | (3.7%) | 1 | (1.7%) | n.s. |
| ST8-MRSA-II | 2 (2.4%) | 1 | (3.7%) | 1 | (1.7%) | n.s. |
| CC8-MRSA-IV | 34 (40.0%) | 6 | (22.2%) | 28 | (48.3%) | 0.032 |
| ST8-MRSA-IV | 32 (37.6%) | 6 | (22.2%) | 26 | (44.8%) | 0.056 |
| ST2516-MRSA-IV | 1 (1.2%) | - | - | 1 | (1.7%) | n.s. |
| ST8465-MRSA-IV | 1 (1.2%) | - | - | 1 | (1.7%) | n.s. |
| CC121-MRSA-V | 2 (2.4%) | - | - | 2 | (3.4%) | n.s. |
| ST121-MRSA-V | 2 (2.4%) | - | - | 2 | (3.4%) | n.s. |

n.s., $p > 0.2$
