## Supplementary material for "Long-Term Impact of Molecular Epidemiology Shifts of Methicillin-resistant *Staphylococcus aureus* on Severity and Mortality of Bloodstream Infection": Table S3

**Supplementary Table 3. Comparison of the patient and strain characteristics according to the major MRSA type**

| | All | ST8-MRSA-IV | ST8-MRSA-I | ST5-MRSA-II | CC1-MRSA-IV | RSA-IV vs ST8-M | | RSAIV vs ST5--I | | RSAIV vs CC1--M | | RSAI vs ST5--N | | RSA-I vs CC1-M | | RSA-II vs CC1-M | |
|---|---|---|---|---|---|---|---|---|---|---|---|---|---|---|---|---|---|
| | (n=85) | (n=32) | (n=19) | (n=16) | (n=8) | *p* value | *q* value | *p* value | *q* value | *p* value | *q* value | *p* value | *q* value | *p* value | *q* value | *p* value | *q* value |
| Patient characteristics | | | | | | | | | | | | | | | | | |
| Age, average | 65.2 ± 2.4 | 67.4 ± 3.9 | 65.6 ± 5.4 | 63.5 ± 5.3 | 68.1 ± 4.3 | n.s. | | 0.163 | | n.s. | | n.s. | | n.s. | | n.s. | |
| Sex, female | 28 (32.9%) | 11 (34.4%) | 4 (21.1%) | 7 (43.8%) | 5 (62.5%) | n.s. | | n.s. | | n.s. | | n.s. | | 0.072 | n.s. | n.s. | |
| Charlson Comorbidity Index | 3.4 ± 0.3 | 3.1 ± 0.5 | 4.8 ± 0.7 | 3.1 ± 0.5 | 3.0 ± 0.8 | n.s. | | n.s. | | n.s. | | n.s. | | n.s. | | n.s. | |
| Hospitalization days when MRSA was detected | 38.5 ± 6.3 | 40.2 ± 8.1 | 32.6 ± 7.0 | 57.8 ± 26.9 | 48.1 ± 12.1 | n.s. | | n.s. | | n.s. | | n.s. | | n.s. | | n.s. | |
| Antimicrobial use within 30 days prior to MRSA detection | 51 (60.0%) | 20 (62.5%) | 9 (47.4%) | 11 (68.8%) | 5 (62.5%) | n.s. | | n.s. | | n.s. | | n.s. | | n.s. | | n.s. | |
| Classification of infection | | | | | | | | | | | | | | | | | |
| Community-acquired | 5 (5.9%) | 2 (6.3%) | 1 (5.3%) | 1 (6.3%) | 0 (0.0%) | n.s. | | n.s. | | n.s. | | n.s. | | n.s. | | n.s. | |
| Healthcare-associated | 14 (16.5%) | 4 (12.5%) | 4 (21.1%) | 2 (12.5%) | 0 (0.0%) | n.s. | | n.s. | | n.s. | | n.s. | | n.s. | | n.s. | |
| Hospital-acquired | 66 (77.6%) | 26 (81.3%) | 14 (73.7%) | 13 (81.3%) | 8 (100%) | n.s. | | n.s. | | n.s. | | n.s. | | n.s. | | n.s. | |
| Source of MRSA infection | | | | | | | | | | | | | | | | | |
| Intravascular device | 35 (41.2%) | 13 (40.6%) | 9 (47.4%) | 7 (43.8%) | 2 (25.0%) | n.s. | | n.s. | | n.s. | | n.s. | | n.s. | | n.s. | |
| Respiratory tract | 4 (4.7%) | 0 (0.0%) | 1 (5.3%) | 3 (18.8%) | 0 (0.0%) | n.s. | | 0.032 | 0.194 | n.s. | | n.s. | | n.s. | | n.s. | |
| Skin/soft tissue or surgical site | 15 (17.6%) | 7 (21.9%) | 2 (10.5%) | 2 (12.5%) | 1 (12.5%) | n.s. | | n.s. | | n.s. | | n.s. | | n.s. | | n.s. | |
| Abdomen | 5 (5.9%) | 2 (6.3%) | 1 (5.3%) | 1 (6.3%) | 1 (12.5%) | n.s. | | n.s. | | n.s. | | n.s. | | n.s. | | n.s. | |
| Bone and joint | 8 (9.4%) | 2 (6.3%) | 3 (15.8%) | 1 (6.3%) | 0 (0.0%) | n.s. | | n.s. | | n.s. | | n.s. | | n.s. | | n.s. | |
| Others | 5 (5.9%) | 1 (3.1%) | 2 (10.5%) | 1 (6.3%) | 1 (12.5%) | n.s. | | n.s. | | n.s. | | n.s. | | n.s. | | n.s. | |
| Unknown | 13 (15.3%) | 7 (21.9%) | 1 (5.3%) | 1 (6.3%) | 3 (37.5%) | n.s. | | n.s. | | n.s. | | n.s. | | 0.065 | n.s. | 0.091 | n.s. |
| SOFA score | 3.4 ± 0.4 | 3.4 ± 0.7 | 3.5 ± 0.9 | 4.4 ± 1.1 | 2.3 ± 0.6 | n.s. | | n.s. | | n.s. | | n.s. | | n.s. | | n.s. | |
| Initial antimicrobial chemotherapy against MRSA | | | | | | | | | | | | | | | | | |
| Vancomycin | 55 (64.7%) | 22 (68.8%) | 12 (62.3%) | 10 (62.5%) | 5 (62.5%) | n.s. | | n.s. | | n.s. | | n.s. | | n.s. | | n.s. | |
| Teicoplanin | 6 (7.1%) | 2 (6.3%) | 1 (5.3%) | 1 (6.3%) | 1 (12.5%) | n.s. | | n.s. | | n.s. | | n.s. | | n.s. | | n.s. | |
| Linezolid | 8 (9.4%) | 3 (9.4%) | 3 (15.8%) | 2 (12.5%) | 0 (0.0%) | n.s. | | n.s. | | n.s. | | n.s. | | n.s. | | n.s. | |
| Daptomycin | 15 (17.6%) | 6 (18.8%) | 3 (15.8%) | 3 (18.8%) | 1 (12.5%) | n.s. | | n.s. | | n.s. | | n.s. | | n.s. | | n.s. | |
| No anti-MRSA agents | 3 (3.5%) | 0 (0.0%) | 1 (5.3%) | 0 (0.0%) | 1 (12.5%) | n.s. | | n.s. | | n.s. | | n.s. | | n.s. | | n.s. | |
| Change of initial treatment | 29 (34.1%) | 14 (43.8%) | 6 (31.6%) | 6 (37.5%) | 2 (25.0%) | n.s. | | n.s. | | n.s. | | n.s. | | n.s. | | n.s. | |
| In-hospital mortality | 19 (22.4%) | 7 (21.9%) | 5 (26.3%) | 5 (31.3%) | 1 (12.5%) | n.s. | | n.s. | | n.s. | | n.s. | | n.s. | | n.s. | |
| 30-days mortality | 11 (12.9%) | 4 (12.5%) | 2 (10.5%) | 4 (25.0%) | 1 (12.5%) | n.s. | | n.s. | | n.s. | | n.s. | | n.s. | | n.s. | |
| Strain characteristics | | | | | | | | | | | | | | | | | |
| Drug resistance rate according to CLSI | | | | | | | | | | | | | | | | | |
| Oxacillin | 81 (95.3%) | 31 (96.9%) | 19 (100%) | 16 (100%) | 7 (87.5%) | n.s. | | n.s. | | n.s. | | n.s. | | n.s. | | n.s. | |
| Cefoxitin | 84 (98.8%) | 32 (100%) | 19 (100%) | 16 (100%) | 8 (100%) | n.s. | | n.s. | | n.s. | | n.s. | | n.s. | | n.s. | |
| Levofloxacin | 65 (76.5%) | 17 (53.1%) | 19 (100%) | 16 (100%) | 8 (100%) | <0.001 | 0.002 | <0.001 | 0.002 | 0.016 | 0.032 | n.s. | | n.s. | | n.s. | |
| Erythromycin | 74 (87.1%) | 22 (68.8%) | 19 (100%) | 16 (100%) | 7 (87.5%) | 0.008 | 0.049 | 0.020 | 0.059 | n.s. | | n.s. | | n.s. | | n.s. | |
| Clindamycin | 36 (42.4%) | 9 (28.1%) | 7 (36.8%) | 16 (100%) | 2 (25.0%) | n.s. | | <0.001 | <0.001 | n.s. | | <0.001 | <0.001 | n.s. | | <0.001 | <0.001 |
| Minocycline | 0 (0.0%) | 0 (0.0%) | 0 (0.0%) | 0 (0.0%) | 0 (0.0%) | n.s. | | n.s. | | n.s. | | n.s. | | n.s. | | n.s. | |
| Vancomycin | 0 (0.0%) | 0 (0.0%) | 0 (0.0%) | 0 (0.0%) | 0 (0.0%) | n.s. | | n.s. | | n.s. | | n.s. | | n.s. | | n.s. | |
| Teicoplanin | 0 (0.0%) | 0 (0.0%) | 0 (0.0%) | 0 (0.0%) | 0 (0.0%) | n.s. | | n.s. | | n.s. | | n.s. | | n.s. | | n.s. | |
| Linezolid | 2 (2.4%) | 0 (0.0%) | 1 (5.3%) | 1 (6.3%) | 0 (0.0%) | n.s. | | n.s. | | n.s. | | n.s. | | n.s. | | n.s. | |
| Drug resistance rate according to EUCAST | | | | | | | | | | | | | | | | | |
| Oxacillin | 81 (95.3%) | 31 (96.9%) | 19 (100%) | 16 (100%) | 7 (87.5%) | n.s. | | n.s. | | n.s. | | n.s. | | n.s. | | n.s. | |
| Cefoxitin | 84 (98.8%) | 32 (100%) | 19 (100%) | 16 (100%) | 8 (100%) | n.s. | | n.s. | | n.s. | | n.s. | | n.s. | | n.s. | |
| Levofloxacin | 65 (76.5%) | 17 (53.1%) | 19 (100%) | 16 (100%) | 8 (100%) | <0.001 | 0.002 | <0.001 | 0.002 | 0.016 | 0.032 | n.s. | | n.s. | | n.s. | |
| Erythromycin | 75 (88.2%) | 23 (71.9%) | 19 (100%) | 16 (100%) | 7 (87.5%) | 0.018 | 0.110 | 0.020 | 0.061 | n.s. | | n.s. | | n.s. | | n.s. | |
| Clindamycin | 37 (43.5%) | 10 (31.3%) | 7 (36.8%) | 16 (100%) | 2 (25.0%) | n.s. | | <0.001 | <0.001 | n.s. | | <0.001 | <0.001 | n.s. | | <0.001 | <0.001 |
| Minocycline | 37 (43.5%) | 7 (21.9%) | 17 (89.5%) | 11 (68.8%) | 1 (12.5%) | <0.001 | <0.001 | 0.004 | 0.007 | n.s. | | n.s. | | <0.001 | <0.001 | 0.027 | 0.041 |
| Vancomycin | 1 (1.2%) | 0 (0.0%) | 0 (0.0%) | 0 (0.0%) | 0 (0.0%) | n.s. | | n.s. | | n.s. | | n.s. | | n.s. | | n.s. | |
| Teicoplanin | 0 (0.0%) | 0 (0.0%) | 0 (0.0%) | 0 (0.0%) | 0 (0.0%) | n.s. | | n.s. | | n.s. | | n.s. | | n.s. | | n.s. | |
| Linezolid | 2 (2.4%) | 0 (0.0%) | 1 (5.3%) | 1 (6.3%) | 0 (0.0%) | n.s. | | n.s. | | n.s. | | n.s. | | n.s. | | n.s. | |
| Aminoglycoside-resistance genes | 82 (96.5%) | 30 (93.8%) | 19 (100%) | 16 (100%) | 7 (87.5%) | n.s. | | n.s. | | n.s. | | n.s. | | n.s. | | n.s. | |
| *aac(6')-aph(2'')* | 44 (51.8%) | 21 (65.6%) | 14 (73.7%) | 3 (18.8%) | 1 (12.5%) | n.s. | | 0.005 | 0.015 | 0.014 | 0.021 | 0.002 | 0.012 | 0.009 | 0.017 | n.s. | |
| *aadD* | 46 (54.1%) | 15 (46.9%) | 18 (94.7%) | 9 (56.3%) | 0 (0.0%) | <0.001 | 0.002 | n.s. | | 0.016 | 0.019 | 0.013 | 0.020 | <0.001 | <0.001 | 0.010 | 0.019 |
| *ant(9)-Ia* | 64 (75.3%) | 15 (46.9%) | 19 (100%) | 16 (100%) | 7 (87.5%) | <0.001 | <0.001 | <0.001 | <0.001 | 0.054 | 0.107 | n.s. | | n.s. | | n.s. | |
| *aph(2')-Ia* | 1 (1.2%) | 0 (0.0%) | 1 (5.3%) | 0 (0.0%) | 0 (0.0%) | n.s. | | n.s. | | n.s. | | n.s. | | n.s. | | n.s. | |
| Beta-lactamase, blaZ | 79 (92.9%) | 29 (90.6%) | 18 (94.7%) | 15 (93.8%) | 8 (100%) | n.s. | | n.s. | | n.s. | | n.s. | | n.s. | | n.s. | |
| Chloramphenicol-resistance genes, *cat(pC221)* | 2 (2.4%) | 1 (3.1%) | 0 (0.0%) | 1 (6.3%) | 0 (0.0%) | n.s. | | n.s. | | n.s. | | n.s. | | n.s. | | n.s. | |
| Fosmycin-resistance | 4 (4.7%) | 0 (0.0%) | 0 (0.0%) | 3 (18.8%) | 0 (0.0%) | n.s. | | 0.032 | 0.194 | n.s. | | 0.086 | n.s. | n.s. | | n.s. | |
| *fosB6* | 3 (3.5%) | 0 (0.0%) | 0 (0.0%) | 3 (18.8%) | 0 (0.0%) | n.s. | | 0.032 | 0.194 | n.s. | | 0.086 | n.s. | n.s. | | n.s. | |
| *fosD* | 1 (1.2%) | 0 (0.0%) | 0 (0.0%) | 0 (0.0%) | 0 (0.0%) | n.s. | | n.s. | | n.s. | | n.s. | | n.s. | | n.s. | |
| Macrolide-resistance | 75 (88.2%) | 23 (71.9%) | 19 (100%) | 16 (100%) | 7 (87.5%) | 0.018 | 0.110 | 0.020 | 0.061 | n.s. | | n.s. | | n.s. | | n.s. | |
| *erm(A)* | 64 (75.3%) | 15 (46.9%) | 19 (100%) | 16 (100%) | 7 (87.5%) | <0.001 | <0.001 | <0.001 | <0.001 | 0.054 | 0.107 | n.s. | | n.s. | | n.s. | |
| *erm(C)* | 13 (15.3%) | 7 (21.9%) | 0 (0.0%) | 2 (12.5%) | 1 (12.5%) | 0.037 | n.s. | n.s. | | n.s. | | n.s. | | n.s. | | n.s. | |
| *msr(A)* | 1 (1.2%) | 1 (3.1%) | 0 (0.0%) | 0 (0.0%) | 0 (0.0%) | n.s. | | n.s. | | n.s. | | n.s. | | n.s. | | n.s. | |
| Tetracycline-resistance | 37 (43.5%) | 10 (31.3%) | 14 (73.7%) | 11 (68.8%) | 1 (12.5%) | 0.004 | 0.026 | 0.029 | 0.044 | n.s. | | n.s. | | 0.009 | 0.026 | 0.027 | 0.054 |
| *Tet (K)* | 2 (2.4%) | 2 (6.3%) | 0 (0.0%) | 0 (0.0%) | 0 (0.0%) | n.s. | | n.s. | | n.s. | | n.s. | | n.s. | | n.s. | |
| *Tet (M)* | 35 (41.2%) | 8 (25.0)% | 14 (73.7%) | 11 (68.8%) | 1 (12.5%) | 0.001 | 0.007 | 0.005 | 0.016 | n.s. | | n.s. | | 0.009 | 0.017 | 0.027 | 0.041 |
| Bleomycin-resistance, *bleO* | 44 (51.8%) | 15 (46.9%) | 17 (89.5%) | 9 (56.3%) | 0 (0.0%) | 0.003 | 0.008 | n.s. | | 0.016 | 0.024 | 0.050 | 0.060 | <0.001 | <0.001 | 0.010 | 0.019 |
| Exoenzyme genes | | | | | | | | | | | | | | | | | |
| *aur* | 85 (100%) | 32 (100%) | 19 (100%) | 16 (100%) | 8 (100%) | n.s. | | n.s. | | n.s. | | n.s. | | n.s. | | n.s. | |
| *splA* | 85 (100%) | 32 (100%) | 19 (100%) | 16 (100%) | 8 (100%) | n.s. | | n.s. | | n.s. | | n.s. | | n.s. | | n.s. | |
| *splB* | 85 (100%) | 32 (100%) | 19 (100%) | 16 (100%) | 8 (100%) | n.s. | | n.s. | | n.s. | | n.s. | | n.s. | | n.s. | |
| *splE* | 43 (50.6%) | 15 (46.9%) | 19 (100%) | 0 (0.0%) | 7 (87.5%) | <0.001 | <0.001 | <0.001 | <0.001 | 0.054 | 0.054 | <0.001 | <0.001 | <0.001 | <0.001 | <0.001 | <0.001 |
| Toxin genes | | | | | | | | | | | | | | | | | |
| *edinA* | 11 (12.9%) | 7 (21.9%) | 0 (0.0%) | 0 (0.0%) | 0 (0.0%) | 0.037 | n.s. | 0.079 | n.s. | n.s. | | n.s. | | n.s. | | n.s. | |
| *eta* | 1 (1.2%) | 0 (0.0%) | 0 (0.0%) | 0 (0.0%) | 0 (0.0%) | n.s. | | n.s. | | n.s. | | n.s. | | n.s. | | n.s. | |
| *hlgA* | 85 (100%) | 32 (100%) | 19 (100%) | 16 (100%) | 0 (0.0%) | n.s. | | n.s. | | n.s. | | n.s. | | n.s. | | n.s. | |
| *hlgB* | 85 (100%) | 32 (100%) | 19 (100%) | 16 (100%) | 8 (100%) | n.s. | | n.s. | | n.s. | | n.s. | | n.s. | | n.s. | |
| *hlgC* | 85 (100%) | 32 (100%) | 19 (100%) | 16 (100%) | 8 (100%) | n.s. | | n.s. | | n.s. | | n.s. | | n.s. | | n.s. | |
| *LukD* | 85 (100%) | 32 (100%) | 19 (100%) | 16 (100%) | 8 (100%) | n.s. | | n.s. | | n.s. | | n.s. | | n.s. | | n.s. | |
| *LukE* | 84 (98.8%) | 32 (100%) | 19 (100%) | 15 (93.8%) | 8 (100%) | n.s. | | n.s. | | n.s. | | n.s. | | n.s. | | n.s. | |
| *LukF-PV* | 2 (2.4%) | 2 (6.3%) | 0 (0.0%) | 0 (0.0%) | 0 (0.0%) | n.s. | | n.s. | | n.s. | | n.s. | | n.s. | | n.s. | |
| *sea* | 11 (12.9%) | 0 (0.0%) | 0 (0.0%) | 0 (0.0%) | 8 (100%) | n.s. | | n.s. | | <0.001 | <0.001 | n.s. | | <0.001 | <0.001 | <0.001 | <0.001 |
| *seb* | 2 (2.4%) | 0 (0.0%) | 0 (0.0%) | 0 (0.0%) | 0 (0.0%) | n.s. | | n.s. | | n.s. | | n.s. | | n.s. | | n.s. | |
| *sec* | 30 (35.3%) | 14 (43.8%) | 0 (0.0%) | 14 (87.5%) | 0 (0.0%) | <0.001 | 0.001 | 0.005 | 0.008 | 0.034 | 0.041 | <0.001 | <0.001 | n.s. | | <0.001 | <0.001 |
| *sec3* | 1 (1.2%) | 1 (3.1%) | 0 (0.0%) | 0 (0.0%) | 0 (0.0%) | n.s. | | n.s. | | n.s. | | n.s. | | n.s. | | n.s. | |
| *sed* | 9 (10.6%) | 1 (3.1%) | 8 (42.1%) | 0 (0.0%) | 0 (0.0%) | <0.001 | 0.005 | n.s. | | n.s. | | 0.004 | 0.011 | 0.061 | 0.121 | n.s. | |
| *seg* | 22 (25.9%) | 0 (0.0%) | 0 (0.0%) | 16 (100%) | 0 (0.0%) | n.s. | | <0.001 | <0.001 | n.s. | | <0.001 | <0.001 | n.s. | | <0.001 | <0.001 |
| *seh* | 9 (10.6%) | 0 (0.0%) | 0 (0.0%) | 1 (6.3%) | 8 (100%) | n.s. | | n.s. | | <0.001 | <0.001 | n.s. | | <0.001 | <0.001 | <0.001 | <0.001 |
| *sei* | 23 (27.1%) | 1 (3.1%) | 0 (0.0%) | 16 (100%) | 0 (0.0%) | n.s. | | <0.001 | <0.001 | n.s. | | <0.001 | <0.001 | n.s. | | <0.001 | <0.001 |
| *sej* | 10 (11.8%) | 1 (3.1%) | 9 (47.4%) | 0 (0.0%) | 0 (0.0%) | <0.001 | 0.001 | n.s. | | n.s. | | 0.002 | 0.005 | 0.026 | 0.052 | n.s. | |
| *sek* | 10 (11.8%) | 1 (3.1%) | 0 (0.0%) | 0 (0.0%) | 8 (100%) | n.s. | | n.s. | | <0.001 | 0.001 | n.s. | | <0.001 | 0.001 | <0.001 | 0.001 |
| *sel* | 29 (34.1%) | 13 (40.6%) | 0 (0.0%) | 13 (81.3%) | 0 (0.0%) | <0.001 | 0.002 | 0.013 | 0.020 | 0.037 | 0.045 | <0.001 | <0.001 | n.s. | | <0.001 | <0.001 |
| *sem* | 23 (27.1%) | 1 (3.1%) | 0 (0.0%) | 16 (100%) | 0 (0.0%) | n.s. | | <0.001 | <0.001 | n.s. | | <0.001 | <0.001 | n.s. | | <0.001 | <0.001 |
| *sen* | 23 (27.1%) | 1 (3.1%) | 0 (0.0%) | 16 (100%) | 0 (0.0%) | n.s. | | <0.001 | <0.001 | n.s. | | <0.001 | <0.001 | n.s. | | <0.001 | <0.001 |
| *seo* | 22 (25.9%) | 0 (0.0%) | 0 (0.0%) | 16 (100%) | 0 (0.0%) | n.s. | | <0.001 | <0.001 | n.s. | | <0.001 | <0.001 | n.s. | | <0.001 | <0.001 |
| *sep* | 24 (28.2%) | 14 (43.8%) | 1 (5.3%) | 9 (56.3%) | 0 (0.0%) | 0.039 | 0.059 | n.s. | | 0.034 | 0.068 | 0.002 | 0.010 | n.s. | | 0.010 | 0.029 |
| *seq* | 10 (11.8%) | 1 (3.1%) | 0 (0.0%) | 0 (0.0%) | 8 (100%) | n.s. | | n.s. | | <0.001 | 0.001 | n.s. | | <0.001 | 0.001 | <0.001 | 0.001 |
| *ser* | 10 (11.8%) | 1 (3.1%) | 9 (47.4%) | 0 (0.0%) | 0 (0.0%) | <0.001 | 0.001 | n.s. | | n.s. | | 0.002 | 0.005 | 0.026 | 0.052 | n.s. | |
| *seu* | 22 (25.9%) | 0 (0.0%) | 0 (0.0%) | 16 (100%) | 0 (0.0%) | n.s. | | <0.001 | <0.001 | n.s. | | <0.001 | <0.001 | n.s. | | <0.001 | <0.001 |
| *tst* | 31 (36.5%) | 14 (43.8%) | 0 (0.0%) | 15 (93.8%) | 0 (0.0%) | <0.001 | 0.001 | 0.001 | 0.002 | 0.034 | 0.041 | <0.001 | <0.001 | n.s. | | <0.001 | <0.001 |
| Others | | | | | | | | | | | | | | | | | |
| ACME | 2 (2.4%) | 2 (6.3%) | 0 (0.0%) | 0 (0.0%) | 0 (0.0%) | n.s. | | n.s. | | n.s. | | n.s. | | n.s. | | n.s. | |
| *sak* | 83 (97.6%) | 30 (93.8%) | 19 (100%) | 16 (100%) | 8 (100%) | n.s. | | n.s. | | n.s. | | n.s. | | n.s. | | n.s. | |
| *scn* | 83 (97.6%) | 30 (93.8%) | 19 (100%) | 16 (100%) | 8 (100%) | n.s. | | n.s. | | n.s. | | n.s. | | n.s. | | n.s. | |

n.s., *P* or *Q values* > 0.2
