## Supplementary material for "Long-Term Impact of Molecular Epidemiology Shifts of Methicillin-resistant *Staphylococcus aureus* on Severity and Mortality of Bloodstream Infection": Table S4

**Supplementary Table 4. Comparison of patient and strain characteristics according to the sug-groups in ST8-IV**

| | CA-MRSA/J (n=13) | t5071-ST8-IV (n=4) | *p* value |
|---|---|---|---|
| Patient characteristics | | | |
| Age, average | 75.2 ± 3.2 | 58.8 ± 19.4 | 0.175 |
| Sex, female | 5 (38.5%) | 2 (50.0%) | n.s. |
| Charlson comorbidity index | 3.8 ± 0.8 | 2.0 ± 0.7 | n.s. |
| Number of days of hospitalization after MRSA detection | 38.1 ± 11.1 | 27.5 ± 27.5 | n.s. |
| Antimicrobial use during the 30 days before MRSA detection | 7 (53.8%) | 1 (25.0%) | n.s. |
| Classification of infection | | | |
| Community-acquired | 0 (0.0%) | 1 (25.0%) | n.s. |
| Healthcare-associated | 2 (15.4%) | 2 (50.0%) | n.s. |
| Hospital-acquired | 11 (84.6%) | 1 (25.0%) | 0.053 |
| Source of MRSA infection | | | |
| Intravascular device | 5 (38.5%) | 1 (25.0%) | n.s. |
| Skin/soft tissue or surgical site | 3 (23.1%) | 1 (25.0%) | n.s. |
| Abdomen | 1 (7.7%) | 0 (0.0%) | n.s. |
| Bone and joint | 0 (0.0%) | 0 (0.0%) | n.s. |
| Others | 1 (7.7%) | 0 (0.0%) | n.s. |
| Unknown | 3 (23.1%) | 2 (50.0%) | n.s. |
| SOFA score | 4.6 ± 1.3 | 1.0 ± 0.0 | 0.166 |
| Initial antimicrobial chemotherapy against MRSA | | | |
| Vancomycin | 10 (76.9%) | 3 (75.0%) | n.s. |
| Teicoplanin | 1 (7.7%) | 0 (0.0%) | n.s. |
| Linezolid | 1 (7.7%) | 1 (25.0%) | n.s. |
| Daptomycin | 1 (7.7%) | 0 (0.0%) | n.s. |
| No anti-MRSA agents | 0 (0.0%) | 0 (0.0%) | n.s. |
| Change of initial treatment | 7 (53.8%) | 1 (25.0%) | n.s. |
| In-hospital mortality | 3 (23.1%) | 1 (25.0%) | n.s. |
| 30-days mortality | 3 (23.1%) | 1 (25.0%) | n.s. |
| Strain characteristics | | | |
| Drug resistance rate according to CLSI | | | |
| Oxacillin | 13 (100%) | 4 (100%) | n.s. |
| Cefoxitin | 13 (100%) | 4 (100%) | n.s. |
| Levofloxacin | 0 (0.0%) | 4 (100%) | <0.001 |
| Erythromycin | 6 (46.2%) | 4 (100%) | 0.103 |
| Clindamycin | 1 (7.7%) | 3 (75.0%) | 0.022 |
| Minocycline | 0 (0.0%) | 0 (0.0%) | n.s. |
| Vancomycin | 0 (0.0%) | 0 (0.0%) | n.s. |
| Teicoplanin | 0 (0.0%) | 0 (0.0%) | n.s. |
| Linezolid | 0 (0.0%) | 0 (0.0%) | n.s. |
| Drug resistance rate according to EUCAST | | | |
| Oxacillin | 13 (100%) | 4 (100%) | n.s. |
| Cefoxitin | 13 (100%) | 4 (100%) | n.s. |
| Levofloxacin | 0 (0.0%) | 4 (100%) | <0.001 |
| Erythromycin | 6 (46.2%) | 4 (100%) | 0.103 |
| Clindamycin | 1 (7.7%) | 4 (100%) | 0.002 |
| Minocycline | 0 (0.0%) | 2 (50.0%) | 0.044 |
| Vancomycin | 0 (0.0%) | 0 (0.0%) | n.s. |
| Teicoplanin | 0 (0.0%) | 0 (0.0%) | n.s. |
| Linezolid | 0 (0.0%) | 0 (0.0%) | n.s. |
| *spa* type | | | |
| t8 | 1 (7.7%) | 0 (0.0%) | n.s. |
| t24 | 0 (0.0%) | 0 (0.0%) | n.s. |
| t351 | 0 (0.0%) | 0 (0.0%) | n.s. |
| t1767 | 5 (38.5%) | 0 (0.0%) | n.s. |
| t2083 | 1 (7.7%) | 0 (0.0%) | n.s. |
| t2229 | 1 (7.7%) | 0 (0.0%) | n.s. |
| t3286 | 0 (0.0%) | 0 (0.0%) | n.s. |
| t5071 | 0 (0.0%) | 4 (100%) | <0.001 |
| t6127 | 0 (0.0%) | 0 (0.0%) | n.s. |
| t16888 | 0 (0.0%) | 0 (0.0%) | n.s. |
| Unknown | 5 (38.5%) | 0 (0.0%) | n.s. |
| Aminoglycoside-resistance genes | 13 (100%) | 4 (100%) | n.s. |
| *aac(6')-aph(2'')* | 13 (100%) | 0 (0.0%) | <0.001 |
| *aadD* | 11 (84.6%) | 0 (0.0%) | <0.001 |
| *ant(9)-Ia* | 2 (15.4%) | 4 (100%) | 0.006 |
| Beta-lactamase, blaZ | 11 (84.6%) | 4 (100%) | n.s. |
| Chloramphenicol-resistance genes, *cat(pC221)* | 0 (0.0%) | 1 (25.0%) | n.s. |
| Fosmycin-resistance | 0 (0.0%) | 0 (0.0%) | n.s. |
| *fosB6* | 0 (0.0%) | 0 (0.0%) | n.s. |
| *fosD* | 0 (0.0%) | 0 (0.0%) | n.s. |
| Macrolide-resistance | 7 (53.8%) | 4 (100%) | n.s. |
| *erm(A)* | 2 (15.4%) | 4 (100%) | 0.006 |
| *erm(C)* | 5 (38.5%) | 0 (0.0%) | n.s. |
| *msr(A)* | 0 (0.0%) | 0 (0.0%) | n.s. |
| Tetracycline-resistance | 2 (15.4%) | 2 (50.0%) | n.s. |
| *Tet (K)* | 1 (7.7%) | 0 (0.0%) | n.s. |
| *Tet (M)* | 1 (7.7%) | 2 (50.0%) | 0.121 |
| Bleomycin-resistance, *bleO* | 11 (84.6%) | 0 (0.0%) | <0.001 |
| Exoenzyme genes | | | |
| *aur* | 13 (100%) | 4 (100%) | n.s. |
| *splA* | 13 (100%) | 4 (100%) | n.s. |
| *splB* | 13 (100%) | 4 (100%) | n.s. |
| *splE* | 0 (0.0%) | 3 (75.0%) | 0.006 |
| Toxin genes | | | |
| *edinA* | 6 (46.2%) | 0 (0.0%) | n.s. |
| *eta* | 0 (0.0%) | 0 (0.0%) | n.s. |
| *hlgA* | 13 (100%) | 4 (100%) | n.s. |
| *hlgB* | 13 (100%) | 4 (100%) | n.s. |
| *hlgC* | 13 (100%) | 4 (100%) | n.s. |
| *LukD* | 13 (100%) | 4 (100%) | n.s. |
| *LukE* | 13 (100%) | 4 (100%) | n.s. |
| *LukF-PV* | 0 (0.0%) | 0 (0.0%) | n.s. |
| *sea* | 0 (0.0%) | 0 (0.0%) | n.s. |
| *seb* | 0 (0.0%) | 0 (0.0%) | n.s. |
| *sec* | 13 (100%) | 0 (0.0%) | <0.001 |
| *sec3* | 0 (0.0%) | 0 (0.0%) | n.s. |
| *sed* | 1 (7.7%) | 0 (0.0%) | n.s. |
| *seg* | 0 (0.0%) | 0 (0.0%) | n.s. |
| *seh* | 0 (0.0%) | 0 (0.0%) | n.s. |
| *sei* | 0 (0.0%) | 0 (0.0%) | n.s. |
| *sej* | 1 (7.7%) | 0 (0.0%) | n.s. |
| *sek* | 0 (0.0%) | 0 (0.0%) | n.s. |
| *sel* | 12 (92.3%) | 0 (0.0%) | <0.001 |
| *sem* | 0 (0.0%) | 0 (0.0%) | n.s. |
| *sen* | 0 (0.0%) | 0 (0.0%) | n.s. |
| *seo* | 0 (0.0%) | 0 (0.0%) | n.s. |
| *sep* | 3 (23.1%) | 4 (100%) | 0.015 |
| *seq* | 0 (0.0%) | 0 (0.0%) | n.s. |
| *ser* | 1 (7.7%) | 0 (0.0%) | n.s. |
| *seu* | 0 (0.0%) | 0 (0.0%) | n.s. |
| *tst* | 12 (92.3%) | 0 (0.0%) | 0.002 |
| Others | | | |
| ACME | 0 (0.0%) | 0 (0.0%) | n.s. |
| *sak* | 12 (92.3%) | 4 (100%) | n.s. |
| *scn* | 12 (92.3%) | 4 (100%) | n.s. |

n.s., *P* or *Q values* > 0.2
