## Supplementary material for "Long-Term Impact of Molecular Epidemiology Shifts of Methicillin-resistant *Staphylococcus aureus* on Severity and Mortality of Bloodstream Infection": Table S5

Supplementary Table 5.Changes in outcomes from 2003 to 2019 with the same inclusion criteria

| | 2003-2007 (n=83) | | 2008-2011 (n=83) | | 2012-2015 (n=70) | | 2016-2019 (n=94) | | 2003-2007 vs 2008-2011 | | 2003-2007 vs 2012-2015 | | 2003-2007 vs 2016-2019 | | 2008-2011 vs 2012-2015 | | 2008-2011 vs 2016-2019 | | 2012-2015 vs 2016-2019 | |
|---|---|---|---|---|---|---|---|---|---|---|---|---|---|---|---|---|---|---|---|---|
| | n | (%) | n | (%) | n | (%) | n | (%) | *P* value | *Q* value | *P* value | *Q* value | *P* value | *Q* value | *P* value | *Q* value | *P* value | *Q* value | *P* value | *Q* value |
| In-hospital mortality | 33 | (39.8%) | 21 | (25.3%) | 17 | (24.3%) | 16 | (17.0%) | 0.068 | 0.136 | 0.057 | 0.170 | 0.001 | 0.008 | n.s. | | 0.198 | n.s. | n.s. | |
| 30-days mortality | No data | | 15 | (18.1%) | 5 | (15.7%) | 10 | (10.6%) | - | - | - | - | - | - | n.s. | | n.s. | | n.s. | |

n.s., *P* or *Q values* > 0.2
